# Short-term intensive fasting improves red blood cell function and rejuvenates erythropoiesis via regulating MS4A3-CDK2 module

**DOI:** 10.1101/2022.08.18.22278875

**Authors:** Li Xu, Wen Wei, Jiawei Qian, Suping Zhang, Zhicong Jin, Xueqing Wang, Lei Li, Yue Gu, Yaqi Lv, Chen Zhao, Xueqin Gao, Wenwen Bu, Ruizhi Zhang, Xiaoya Ma, Yanjun Yang, Zhanjun Yan, Yuwei Chen, Qiyuan Sun, Li Wang, Zixing Chen, Youjia Xu, Peng Xu, Yun Zhao, Yixuan Fang, Na Yuan, Jianrong Wang

**Author notes:** These authors contributed to this work equally (Li Xu, Wen Wei, Jiawei Qian). Correspondence: Jianrong Wang, Na Yuan, Yixuan Fang.

## Abstract

Fasting is known to improve health, but the precisely beneficial effects of specific types of fasting and their underlying mechanisms are not fully understood. We herein report that in humans, occasional short-term intensive fasting (STIF), a traditional fasting format favored by Asians, promotes erythropoiesis and boosts the function of red blood cells (RBCs) in oxygen transportation, ATP generation, antioxidant capacity, and innate immune response. The rejuvenation of erythropoiesis is more pronounced in humans with low RBC counts. Using mouse models and a human erythroid progenitor cell model, we found that occasional STIF rejuvenates erythropoiesis by enhancing megakaryocyte-erythroid progenitor selfrenewal and erythroid-biased differentiation without compromising normal hematopoiesis. Molecularly, STIF relies on an autophagy-dependent but erythropoietin (EPO) upregulation-independent MS4A3-CDK2 module to augment the production of RBCs. Our findings thus suggest that STIF can occasionally be practiced as an efficient noninvasive intervention for better erythropoiesis, particularly for adults with low RBC counts.

**Graphical abstract:** 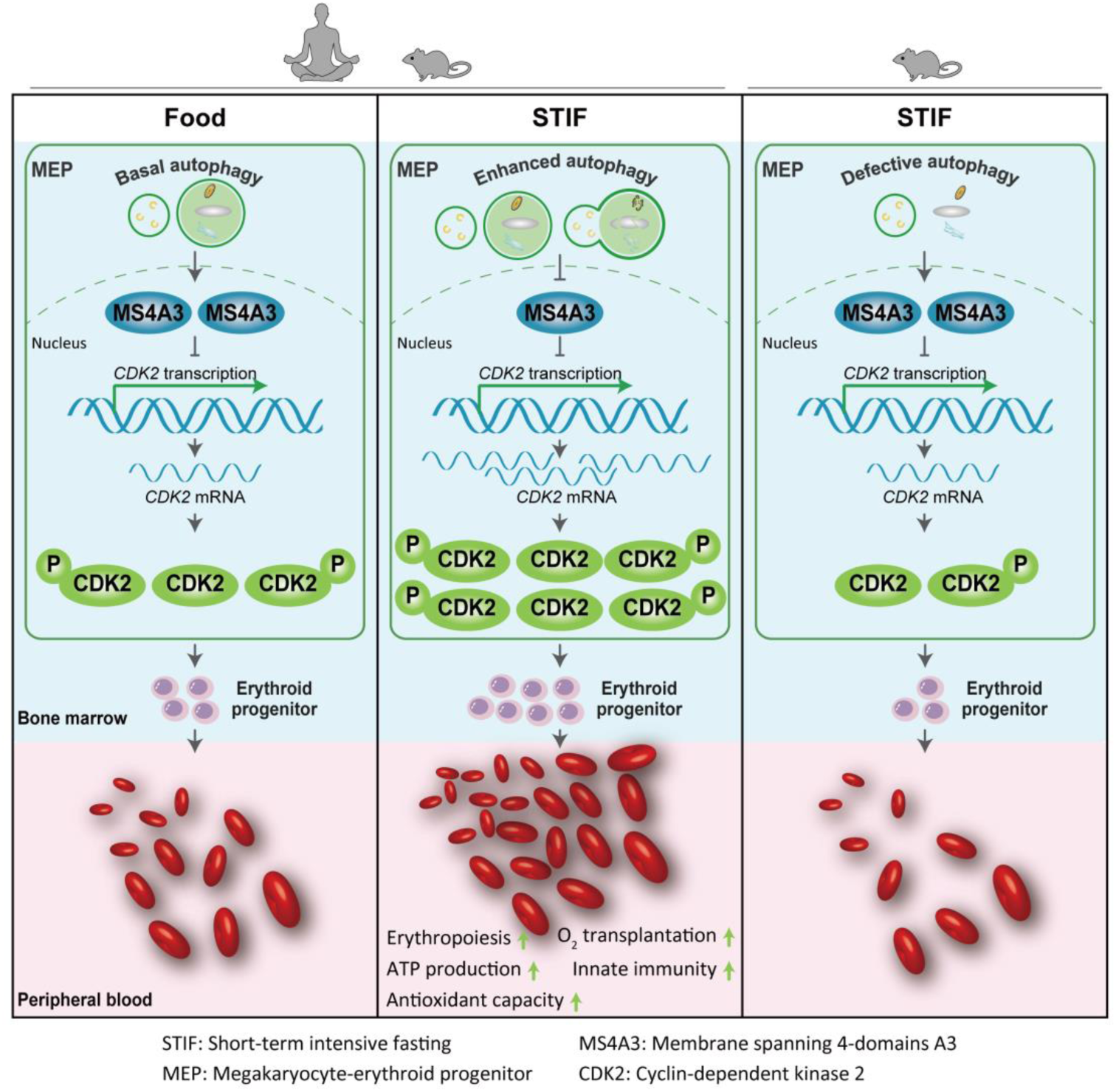

**In brief:** Xu et al showed that short-term intensive fasting (STIF), when occasionally practiced, boosts red blood cell function and promotes erythropoiesis by regulating MS4A3-CDK2 module to enhance megakaryocyte-erythroid progenitor selfrenewal and erythroid-biased differentiation.

**Highlights:**

- Occasional STIF boosts the function of red blood cells
- Occasional STIF promotes erythropoiesis, which is more significant in adults with low red blood cell counts
- Occasional STIF selectively enhances MEP selfrenewal and erythroid-biased differentiation via MS4A3-CDK2 module
- The erythropoietic MS4A3-CDK2 response to STIF is autophagy-dependent but EPO upregulation-independent.

## INTRODUCTION

Low production of red blood cells (RBCs) is one of the most prevalent subhealth conditions in adults worldwide (Kassebaum et al., 2014; Kassebaum, 2016; Jaiswal and Ebert, 2019; Jaiswal, 2020). The capacity to generate RBCs declines with human physiological aging and pathological conditions, such as anemia, myelodysplastic syndrome, leukemia, or other diseases where the body is immune compromised (Sankaran and Weiss, 2015; Stauder et al., 2018; Jaiswal and Ebert, 2019; Jaiswal, 2020). Low RBC count is also associated with a number of other clinically relevant conditions, including cardiovascular diseases (Culleton et al., 2006), cognitive impairment (Denny et al., 2006; Hong et al., 2013), insomnia (Chen-Edinboro et al., 2017), depression (Onder et al., 2005) and reduced physical performance (Penninx et al., 2004; Chaves et al., 2002). In addition to blood transfusion and nutrient supplementation, erythropoiesis-stimulating agents, such as recombinant human erythropoietin (EPO), have been the main font-line therapy for anemia due to chronic kidney disease and cancer patients undergoing chemotherapy (Sankaran and Weiss, 2015). Recently, gene editing has been explored to treat congenital forms of anemia, such as β-Hb disorders (Wu et al., 2019; Doudna, 2020; Frangoul et al., 2021; Fu et al., 2022). However, for the adult population with a chronic low RBC count, there has been no effective medical solution.

Dietary restriction and fasting of various formats are becoming popular interventions for many health conditions (de Cabo and Mattson, 2019; Longo et al., 2021). Lifelong, not short-term caloric restriction, slows hematopoietic aging in mice (Chen et al., 2003). Prolonged intermittent fasting with 6 cycles of chemotherapy lasting for approximately 80 days promotes the regeneration of hematopoietic stem cells (HSCs) and reverses immunosuppression in mice (Cheng et al., 2014). Long-term dietary restriction for 9 months from young mice to midlife mice improves the maintenance of the repopulation capacity of HSCs by enhancing stem cell quiescence but impairs the lymphoid differentiation capacity of hematopoietic stem and progenitor cells, resulting in immune defects in the context of prolonged bacterial infection (Tang et al., 2016). Encouragingly, a regimen consisting of six cycles of intermittent fasting (12 days) selectively inhibits engrafted B-cell acute lymphoblastic leukemia development via leptin receptor upregulation in host mice (Lu et al., 2017), long-term caloric restriction for 42 months delays T cell senescence in aged-long-lived nonhuman primates (Messaoudi et al., 2006), and dietary restriction triggers bone marrow protection and optimization of immunological memory (Collins et al., 2019), and intermittent caloric restriction promotes erythroid development and ameliorates phenylhydrazine-induced anemia in mice (Bai et al., 2022).

However, the impact of short-term intensive fasting (STIF), a traditional fasting practice initiated in ancient China more than two thousand years ago, remains fundamentally unexplored. Here, we investigated whether and how STIF regulates erythropoiesis in humans, together with the use of mouse models and a human erythroid progenitor cell model to comprehend the underlying mechanisms.

## RESULTS

### 1. STIF boosts the function of red blood cells and rejuvenates erythropoiesis in humans

Thirty-one subjects performed STIF for seven days, a duration favored by water-only fasting participants in China (Figure 1A, Table S1). The STIF increased red blood cell (RBC) and hemoglobin (HGB) levels, which reflect the amount of HGB in the blood, and hematocrit (HCT), which reflects the percentage of RBCs in the blood (Figure 1B). Glycophorins A (GPA) is a major sialoglycoprotein of the human erythrocyte membrane and plays an important role in prevention of red cell aggregation in the circulation (Poole 2000). It is also required for the activity of SLC4A1 that functions as a chloride/bicarbonate exchanger involved in carbon dioxide transport from tissues to lungs (Young et al., 2000; Bruce 2004; Shnitsar et al., 2013). GPA^+^RBCs was increased not only after 7-day fasting, but also after 7-day refeeding, suggesting that this RBC population is prone to maintain a relative higher level after fasting (Figure 1C). Blood smears did not show noticeable changes in morphology (Figure 1D), nor did the average size of RBCs, represented by the mean corpuscular volume (MCV), and the range in the volume and size of RBCs, represented by the red cell distribution width (RDW) change (Figure S1A). 2,3-diphosphoglycerate (2,3-DPG) in RBCs regulates oxygen binding to HGB (MacDonald, 1977). Fasting did not alter the level of 2,3-DPG (Figure S1B, left), suggesting no change in oxygen-carrying function per HGB protein. Furthermore, the SpO_2_ assay indicated that fasting did not change the level of oxygenated HGB in the RBCs (Figure S1B, right), reflecting normal oxygen transportation capacity per HGB molecule in the body. Fasting did not alter the apoptosis of RBCs (Figure S1C). The above data suggest that the increase in RBC count results from more production, not less removal by cell death.

**Figure 1.**
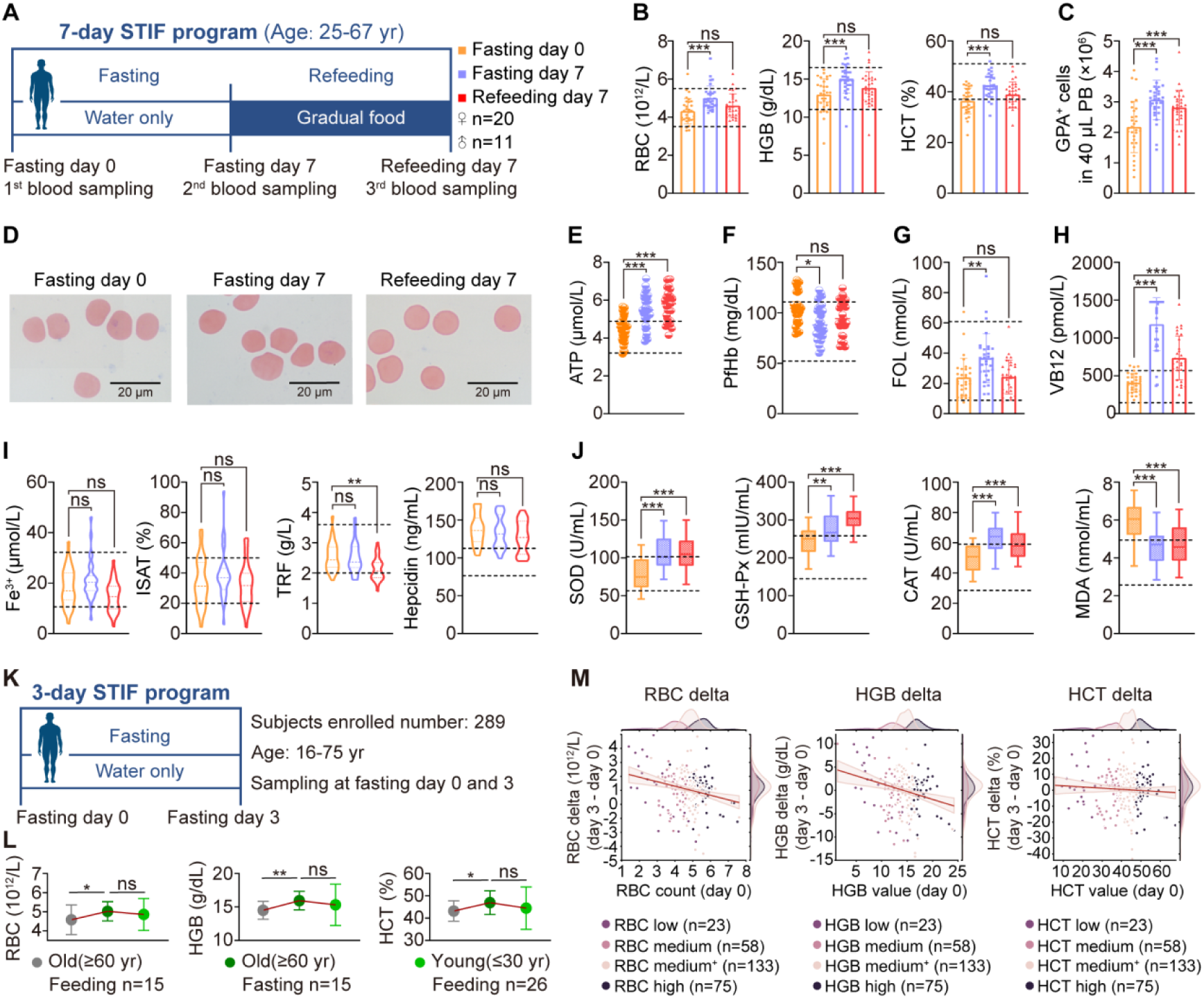
STIF promotes erythropoiesis and improves the function of red blood cells in humans. **(A)** Schematic presentation of the STIF program that includes fasting for 7 days and gradual refeeding for 7 days (n=31). **(B)** Peripheral red blood count. RBC, hemoglobin and HCT by blood routine test. **(C)** Flow cytometric quantitation of human peripheral red blood cells with GPA antibody. **(D)** Peripheral blood smear by Wright-Giemsa stain. **(E)** ATP concentration in peripheral red blood cells detected by ELISA. **(F)** Plasma free hemoglobin (PfHb) measured by ELISA. **(G, H)** The concentrations of Folic acid and vitamin B12 in serum detected by electrogenerated chemiluminescence. **(I)** Serum Fe^3+^ concentration measured by iron colorimetric assay; iron saturation ratio (ISAT) in serum by calculation; transferrin (TRF) in serum by immune transmission turbidity assay; hepcidin in plasma by ELISA. **(J)** SOD, GSH-Px, CAT and MDA of peripheral RBCs tested by ELISA. **(K)** Schematic presentation of the 3-day STIF. **(L)** Peripheral routine RBC count from young participants with normal food and aged participants before and after STIF for 3 days. **(M)** The lower the RBC count before fasting, the greater the increase in the RBC count after STIF. Routine blood data of 289 volunteers for 3-day STIF from multiple independent interventions in different cities were collected. K means was used to divide the RBC, HGB and HCT data before fasting into 4 groups, and the number of RBCs (or HGB, HCT) in these 4 groups ranged from low to high. The abscissa is the RBC (or HGB, HCT) number before fasting (from low to high). The ordinate is the difference between the RBC (or HGB, HCT) numbers after and before 3-day STIF. The line represents the fitted linear regression model and a 95% confidence interval for that regression.

STIF elevated ATP levels in RBCs, and this increase was well sustained after 7 days of refeeding (Figure 1E), suggesting that RBCs retain a better capacity in mechanical properties and chemical signaling processes involved in blood microcirculation, such as shearing stresses (Betz et al., 2009; Zhang et al., 2018). Plasma free hemoglobin (PfHb) causes excessive RBC breakdown and induces vasoconstriction, and increasing PfHb levels is associated with increased vasoconstriction but decreased microvascular density (Damiani et al., 2015). Therefore, a decreased level of PfHb release after fasting (Figure 1F) suggests that RBCs have a reduced risk of damaging endothelial function.

In agreement with the RBC increase, folic acid and vitamin B12, two essential components for helping iron function properly and synthesis of RBCs in the body, were also upregulated in response to fasting (Figure 1G and H). Examination of serum free iron ions, serum iron saturation ratio (ISAT), serum transferrin (TRF) level, and hepcidin did not show alterations by fasting (Figure 1I), indicating that fasting does not disturb iron hemostasis, which is needed in the formation of RBCs. Despite TRF reduction after 7 days of refeeding, free iron ions, ISAT and hepcidin all remained unchanged, suggesting that iron homeostasis was not disturbed by STIF.

Spectrophotometric assays of the principal parameters against oxidative stress indicated that superoxide dismutase (SOD), catalase (CAT) and glutathione peroxidase (GPX) in RBCs all increased, but alondialdehyde (MDA), a product of lipid peroxidation in RBCs, decreased (Figure 1J), suggesting that RBCs have sustainably strengthened power in counteracting oxidative stress after STIF.

Analysis of a hospital record of routine blood tests from physical examination of 4721 people indicates that RBCs decrease in number in the aged population since humans aged more than 60 years have significantly lower RBC counts than young and middle-aged populations (Figure S1D). Shorter term intensive fasting is easier to practice, particularly for aged people. Therefore, we examined the impact of 3-day STIF on erythropoiesis (Figure 1K), and found that the 3-day intervention was sufficient to increase the number of RBCs and the levels of HGB and HCT in the subjects aged more than 60 years to a level comparable to that in the young population aged less than 30 years (Figure 1L).

To confirm the positive outcome of 3-day STIF on erythropoiesis in a larger population, we analyzed 289 volunteers for 3-day STIF from multiple independent interventions in different cities in China, and found that the lower the RBC count before fasting was, the greater the increase in RBC count after fasting (Figure 1M).

### 2. STIF modulates immune response in human red blood cells

We acquired proteomics profiles on RBCs from 18 STIF subjects to identify the difference in protein abundance before and after STIF/refeeding. Differential analysis at the three time points yielded 84 differentially expressed proteins, forming different expression dynamic patterns in the fasting/refeeding process (Figure 2A, Table S2). During the process, 48 proteins were upregulated after STIF, while 17 proteins were downregulated. The abundance of 28 proteins remained unchanged after the refeeding process, while 54 proteins returned to the original abundance. The significantly differentially expressed proteins were enriched in 25 pathways (Table S3). Pathways such as activation of the immune response, proteolysis and response to nitrogen compounds were mostly enriched among these pathways. We used GSEA instead of bare enrichment analysis to incorporate the DEP analysis results into the enrichment process to quantify the positive or negative effect in a given pathway. Despite FDR, GSEA revealed that STIF positively regulates activation of immune response, complement activation and cell activation involved in immune response (Figure 2B), and negatively regulates myeloid leukocyte activation, response to nitrogen compound and organonitrogen compound biosynthetic process (Figure 2B), while refeeding positively regulates proteolysis, complement activation and adaptive immune response (Figure 2C), and negatively regulates organonitrogen compound biosynthetic process, myeloid leukocyte activation (Figure 2C). The immune response of RBCs was activated after STIF (Figure 2D), which was partially reversed after refeeding (Figure 2E).

**Figure 2.**
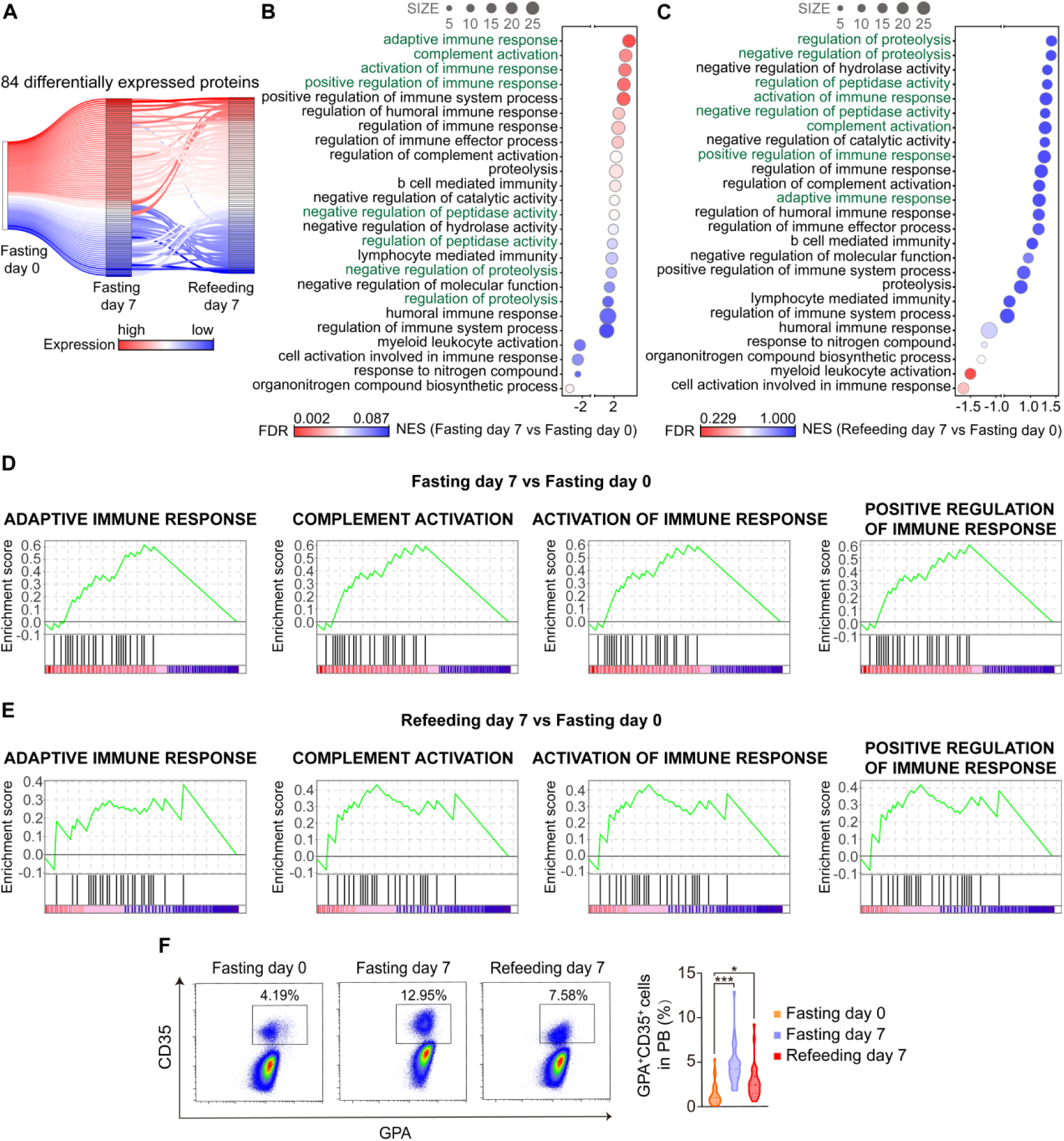
STIF enriches immune response pathways in human red blood cells. **(A)** Expression dynamics of all differentially expressed genes between STIF day 0, STIF day 7 and refeeding day 7. Protein expression at the two time points was normalized to STIF day 0 as relative expression. **(B,C)** Summary of significant gene ontology pathways from GSEA. Color corresponds to false-discovery rate. Size of the dot represents the DEPs in each pathway. The horizontal axis represents the normalized enrichment score from GSEA. **(D,E)** Representative GSEA plots of immune functions. **(F)** Flow cytometric quantitation of the immune function of RBCs with CD35 and GPA antibodies. Fasting day 0 (yellow); fasting day 7 (blue); refeeding day 7 (red), n=31/group.

The expression of CD35 protein on RBCs, which positively correlates with the enhanced immune response in RBCs, was significantly increased after STIF (Figure 2F). Moreover, negative regulation of peptidase activity and negative regulation of proteolysis were increased after STIF (Figure S2A), which was partially reversed after refeeding (Figure S2B). This may partly explain why some proteins increased after STIF. Venn diagram depicts the intersection of DEPs with tighter criteria (adjusted *P* value < 0.05) of two comparison groups (fasting day 7 versus fasting day 0 and refeeding day 7 versus fasting day 0). Twelve proteins were upregulated at STIF day 7 and maintained a high level of expression after refeeding. Twenty-nine proteins were upregulated at STIF day 7 but returned to normal after refeeding. Fourteen proteins were downregulated at refeeding day 7 (Figure S2C). Subsequently, we examined the molecular functions of DEPs from different expression patterns during the fasting/refeeding process and noticed that the main change lies in cell recognition, cell activation, and in particular immune response (Figure S2D-F).

### 3. Occasional, not frequent STIF, gives rise to sustainable increase in red blood cells in mice

To examine whether STIF-enhanced erythropoiesis in humans can be seen in model animals, we fasted 8-week-old mice with water only for 6 hours, which in entire mouse lifespan roughly equals to 3-7 day STIF for entire human lifespan. We observed similar increases in RBC counts, HGB and HCT levels after 6 h fasting (Figure 3A, B). Peripheral RBC counts declined with mouse aging (Figure S3A). 20-month-old mice also showed an increase in RBC counts in response to 6 h fasting (Figure 3C). The functions of bone marrow erythroid cells from old mice after 6 h fasting, measured by reactive oxygen species that reflects oxidative stress, mitochondrial mass that reflects the capacity of generating ATP, and 2,3-DPG that reflects the capacity of HGB carrying oxygen, and the viability of bone marrow erythroid cells measured by plasma free hemoglobin and apoptotic cell death all improved, to the levels of young mice (Figure D-G, S3B). Consistent with the peripheral RBC increase, 6 h fasting increased bone marrow erythroid cells measured by flow cytometry (Figure 3H).

**Figure 3.**
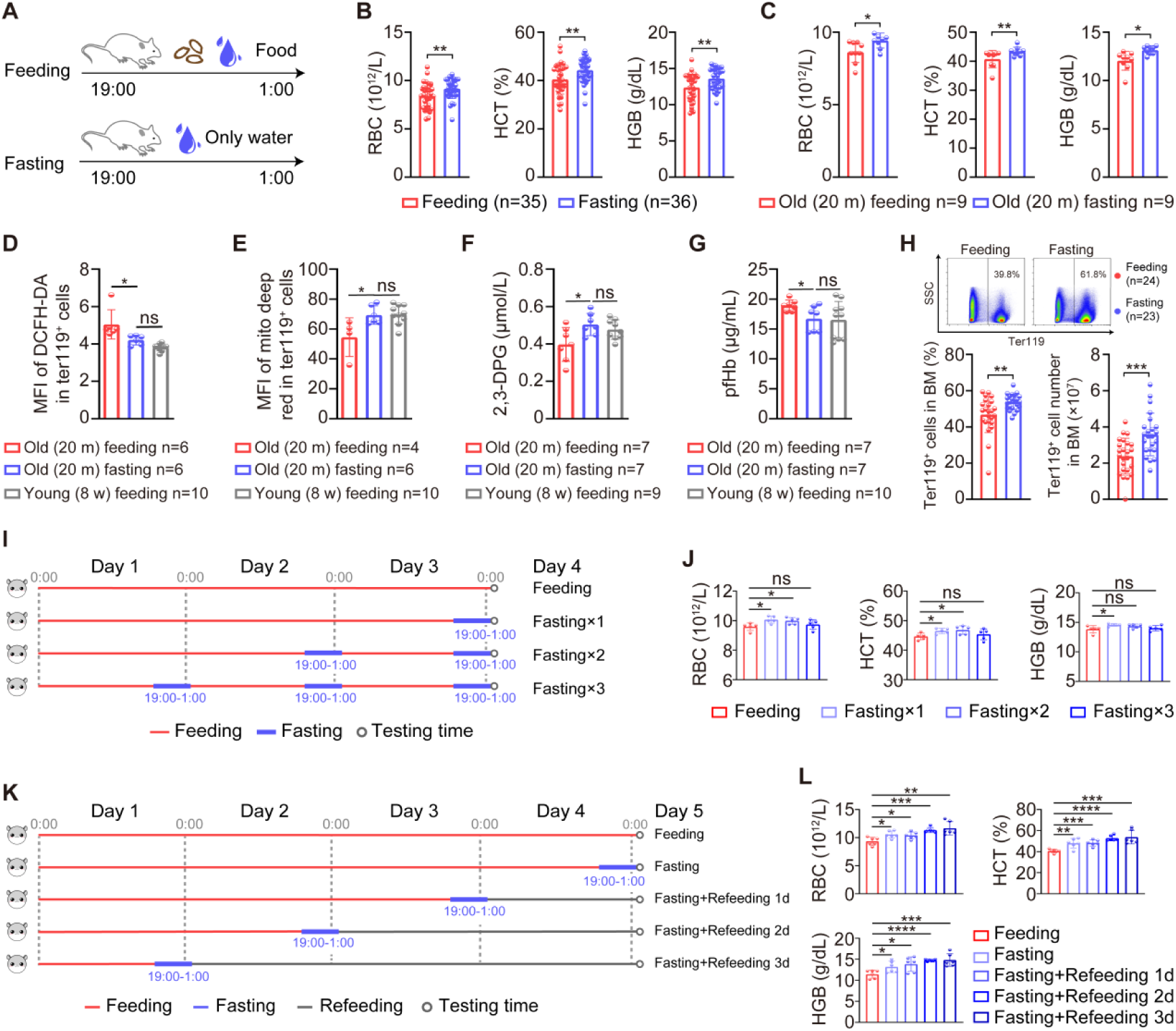
Occasional STIF contributes to the improved function and relatively sustainable increase in red blood cells in mice. **(A)** Schematic program for mouse fasting. **(B)** Peripheral red blood count, HGB and HCT by blood routine test. **(C)** The number of peripheral red blood cell in aged mice before and after fasting was measured by routine blood count. **(D)** ROS levels in mouse bone marrow red blood cells measured with Ter119 and DCFH-DA by polychromatic flow cytometry. **(E)** Mitochondrial mass in mouse bone marrow red blood cells measured with Ter119 and MitoTracker by flow cytometry. **(F)** Oxygen transportation capacity for mouse peripheral red blood cells with a 2,3-DPG ELISA kit. **(G)** Plasma free hemoglobin (pfHb) level in mouse plasma with ELISA kit. **(H)** Flow cytometric quantitation of RBCs with Ter119 antibody. Up, representative gating. Down, statistical numbers for RBCs in the bone marrow. **(I)** Schematic program for once or repeated 6 h fasting. **(J)** RBC, hemoglobin and HCT by blood routine test (n=5/group). (K) Schematic diagram for testing time after 6 h fasting. **(L)** RBC, hemoglobin and HCT by blood routine test (n=6/group).

To determine whether repeated or intermittent STIF is superior to occasional STIF in enhancing RBC generation, we performed 6 h fasting for once, twice or three times in mice (Figure 3I). The results indicated that 6 h fasting once caused increase in RBC count, HGB value and HCT percentage; 6 h fasting twice caused increase in RBC count and HCT percentage, not in HGB value; 6 h fasting three times caused no increase in any of these three parameters of red blood cells (Figure 3J). The results suggest that occasional STIF, not frequent STIF, is able to promote the formation of RBCs.

To answer whether STIF-enhanced erythropoiesis is sustainable, we measured peripheral red blood cell counts of fasting mice in a time course (Figure 3K) and found that after three days of regular feeding following 6 h fasting, red blood cells maintained significantly higher counts with elevated HGB and HCT levels than that before 6 h fasting in mice (Figure 3L). Roughly, three days in mouse lifespan is equivalent to four months in human lifespan. Thus, the result suggests a relatively long-lasting rejuvenation of erythropoiesis by occasional STIF.

### 4. STIF selectively enhances MEP selfrenewal and erythroid-biased differentiation without disturbing HSCs and megakaryocyte progenitors

To understand cellular mechanism by which STIF promotes erythropoiesis in humans, we investigated the progression of erythropoiesis in response to 6 h fasting in mice (Figure 4A). Although fasting does not alter the peripheral red blood cell morphology (Figure S3C), or compromise the proliferation capacity and apoptosis levels of total bone marrow erythroid cells and peripheral RBCs (Figure S3D-F), examination of bone marrow erythroblasts indicates specifically accelerated proliferation of stage IV erythroblasts, namely, orthochromatic erythroblasts (Ter119^hi^CD71^−^), by fasting both at percentage of all bone marrow cells and absolute number in the bone marrow (Figure 4B), whereas spleen erythropoiesis and spleen coefficient were not changed (Figure 4C, S3G), suggesting that erythropoiesis is enhanced by fasting solely in the bone marrow, not extramedullary organ and thus, the fasting-triggered increase in erythropoiesis is a physiological response. Further examination of hematopoietic stem cells (HSCs) and hematopoietic progenitors indicated that fasting specifically increased the pool of MEP cells (Figure 4D), which are the bipotent hematopoietic stem cells responsible for the generation of their downstream magakaryocytes and erythrocytes. Noticeably, increase in MEP pool by STIF was relatively maintainable since MEP number remained high level 3 days after 6 h fasting (Figure S4). Colony formation assays indicated that fasting increased the proliferation and differentiation capacity of GEMM (granulocyte, erythrocyte, macrophage and megakaryocyte) progenitors, and specifically fasting increased the colony formation capacity of erythroid progenitors (CFU-E), not granulocyte and megakaryocyte progenitors (CFU-GM) (Figure 4E). Flow cytometric analysis confirmed that fasting did not increase the percentage or number of megakaryocyte progenitors (Figure 4F). These results indicate that fasting specifically promotes MEP selfrenewal and further erythroid-biased MEP differentiation without causing extramedullary erythropoiesis or altering the generation of megakaryocyte progenitors.

**Figure 4.**
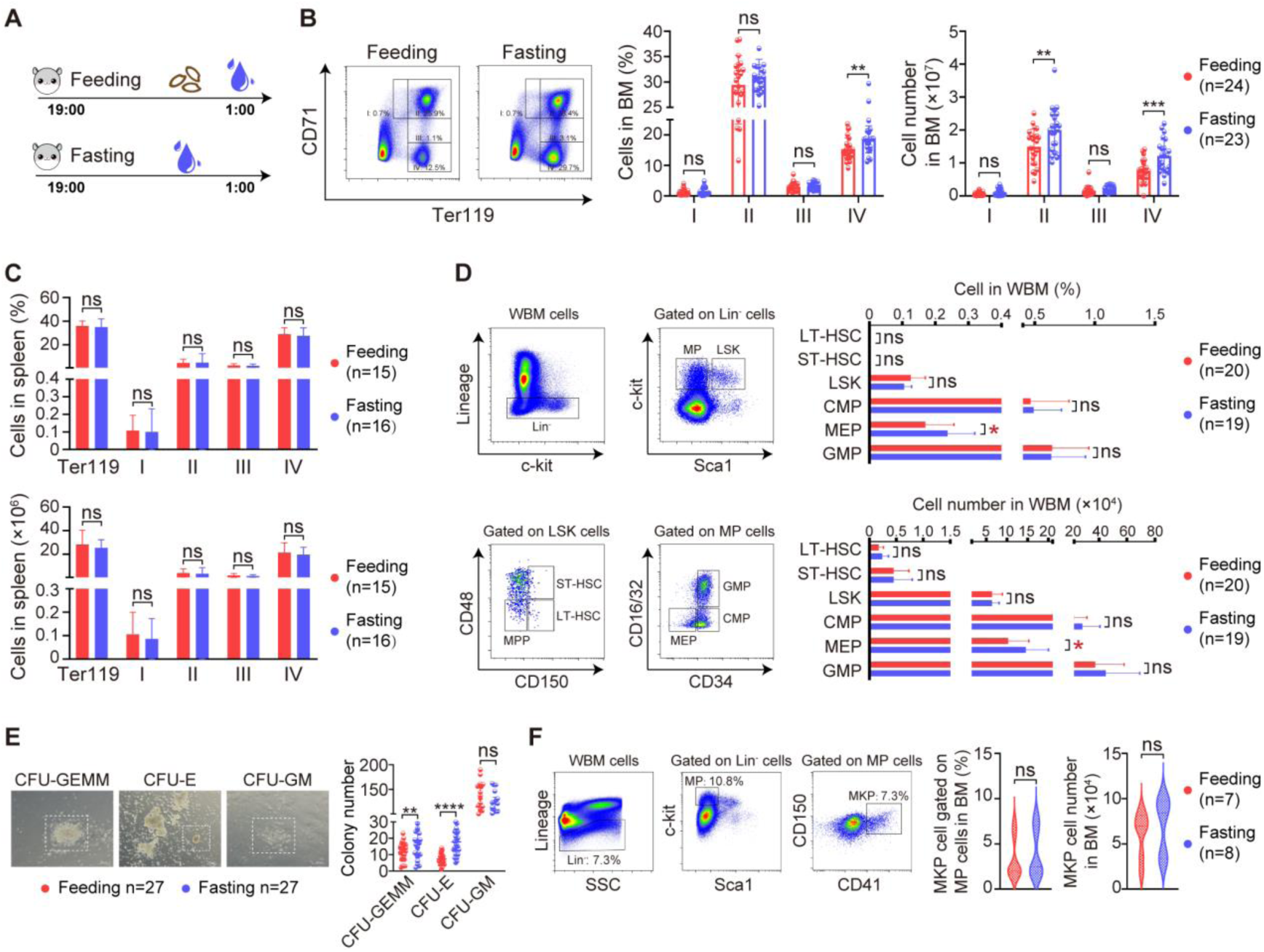
STIF selectively promotes MEP self-renewal and erythroid-biased differentiation without compromising HSPCs and megakaryocyte progenitors in mice. **(A)** Schematic program for mouse fasting. **(B)** Flow cytometric determination of developmental stages in a bone marrow erythrocytic series using fluorescent antibodies against Ter119 and CD71. I: proerythroblast (Ter119^med^CD71^hi^); II: basophilic erythroblast (Ter119^hi^CD71^hi^); III: polychromatophilic erythroblast (Ter119^hi^CD71^med^); IV: orthochromatic erythroblast (Ter119^hi^CD71^−^). Left, representative gating. Right, statistical numbers for erythroid cells in the bone marrow. **(C)** Flow cytometric determination of developmental stages in spleen erythrocytes using fluorescent antibodies against Ter119 and CD71. **(D)** Quantitation of bone marrow stem and progenitor cells on a polychromatic flow cell analyzer. Left, representative gating. Right panel, statistical numbers for hematopoietic stem and progenitor cells. **(E)** Bone marrow progenitor colony assay. Left, colony morphology. Right, statistic results of the colonies. **(F)** Quantitation of bone marrow megakaryocyte progenitors on a polychromatic flow cell analyzer. Left, representative gating. Right, statistical results for megakaryocyte progenitors.

### 5. *Ms4a3* is the most transcriptionally downregulated cell-cycle regulatory gene in cycling-accelerated MEPs in response to STIF

To uncover the molecular signatures involved in the increase of MEP cells in response to STIF, we conducted transcriptomic profiling on MEP (Lin^−^Sca-1^−^c-kit^+^CD34^−^CD16/32^−^) cells from the mice fasted for 6 hours. Principal component analysis showed distinct expression profiles between nonfasting and fasting mice (Figure 5A). Differential analysis of the profile yielded a total of 2425 differentially expressed genes, which consisted of 339 upregulated genes and 2086 downregulated genes (Figure 5B). Enrichment analysis of the DEGs revealed a series of functions that were influenced after fasting, among which the cell cycle process was enhanced (Figure 5C). Significant genes with altered expression levels involved in the cell cycle process are plotted in Figure 5D. We found that one particular gene, *Ms4a3*, demonstrated the highest fold change ratio. The *Ms4a3* gene encodes a member of the membrane-spanning 4A gene family. It is a hematopoietic modulator of the G1-S cell cycle transition upstream of CDK2 (Donato et al., 2002). The downregulation of *Ms4a3* expression in MEP cells in response to 6 h fasting was validated with quantitative PCR (Figure 5E).

**Figure 5.**
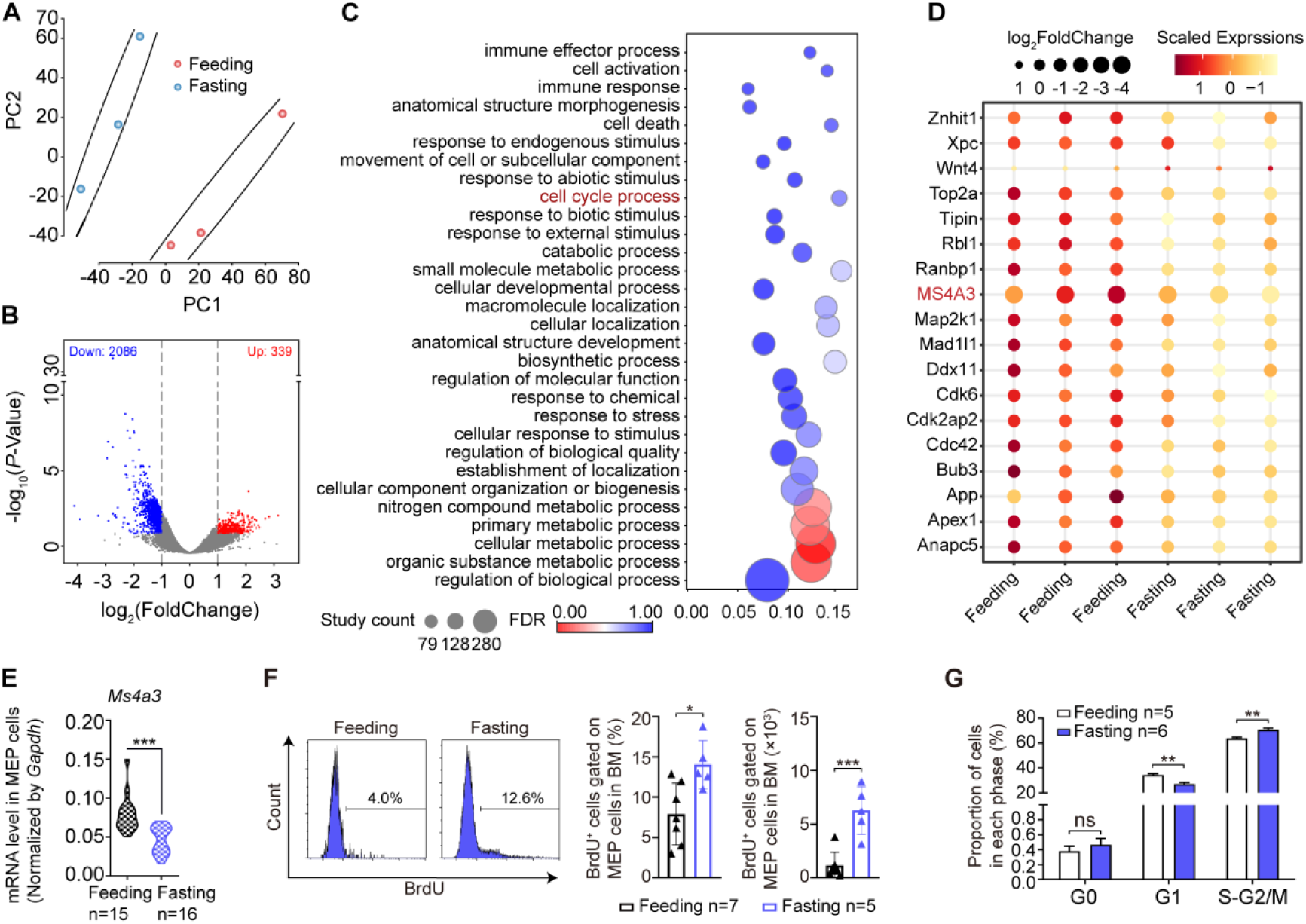
Transcriptomic profiling of MEPs reveals *Ms4a3* as a major downregulated gene in response to STIF in mice. **(A)** Two-dimensional PCA of the MEP cell transcriptome was performed on samples before and 6 h after water-only fasting (n=6). **(B)** Volcano plot of MEP cell transcriptome. In the volcano plot, differentially expressed genes were determined under the condition of fold change≥2 and false discovery rate (adjusted *P* value) threshold≤05. The fold change was log-transformed. Negative values represent a decrease in gene expression, and positive values represent an increase in gene expression. The *P* value was transformed by −log_10_. Red dots represent upregulated genes, and blue dots represent downregulated genes (n=6). **(C)** Gene ontology enrichment analysis of biological processes. Dot size represents the number of genes in each pathway, and the color ranging from red to blue corresponds to statistical significance, FDR, from most significant to least significant (n=6). **(D)** Heatmap of cell cycle negative regulators. **(E)** *Ms4a3* gene expression in MEPs measured by quantitative PCR. **(F)** Measurement of MEP cell proliferation by BrdU staining. **(G)** Measurement of the MEP cell cycle with Ki67-AF700 and Hoechst-33342 double fluorescent staining on a polychromatic flow cell analyzer.

Consistent with transcriptional upregulation of cell cycle positive regulators and downregulation of negative regulators disclosed by transcriptomic analysis, a cell proliferation assay with BrdU incorporation indicated that 6 h fasting speeds up MEP self-renewal or cycling (Figure 5F) without alteration of its quiescence (Figure 5G).

### 6. STIF promotes erythropoiesis by regulating the MS4A3-CDK2 module

Transcriptomic analysis of mouse MEPs indicated that *Ms4a3* mRNA reduction by fasting, alone with upregulation of MEP selfrenewal/proliferation and differentiation, led to increased RBCs. To test whether downregulation of *MS4A3* is required to promote the maturation of RBCs in humans, we applied HUDEP-2, a human cord HSC-derived erythroid progenitor cell line that can be induced to differentiate into mature RBCs (Kurita et al., 2013). *MS4A3* in the HUDEPs was silenced with the *MS4A3* lentiviral vector, which was tested for sufficient viral packing capacity with a GFP marker (Figure 6A). Flow cytometry and western blotting confirmed the successful knockdown of *MS4A3* in human HUDEPs (Figure 6B, C). After silencing *MS4A3*, the HUDEP cells produced more messengers of α, β and γ-globins needed for the synthesis of hemoglobin (Figure 6D) and more mature RBC surface marker GPA in Hoechst negative cells, suggesting more mature RBCs produced by knockdown of *MS4A3* (Figure 6E).

**Figure 6.**
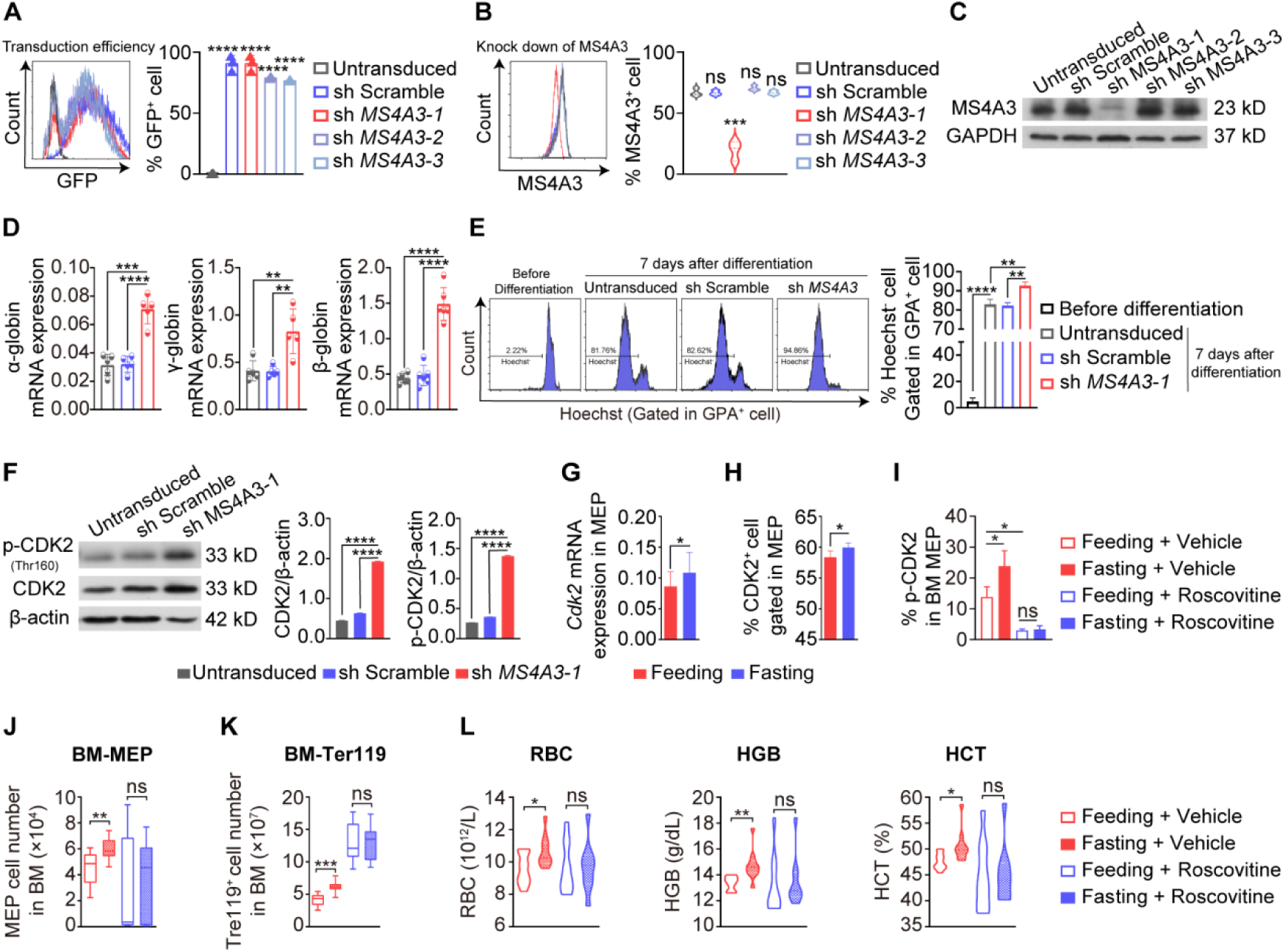
STIF regulates the MS4A3-CDK2 module to promote erythropoiesis. **(A)** HUDEP-2 cells were transduced with a nontargeting scramble shRNA (SCR) and *MS4A3* shRNA. Transduction efficiency was measured 72 h posttransduction by flow cytometry (n=3/group). **(B,C)** Protein level of MS4A3 measured by flow cytometry and western blot following knockdown of MS4A3 by shRNA. A nontargeting scramble shRNA (SCR) was included as a control. Untransduced cells (gray), SCR (blue), *MS4A3* shRNA (red) (n=3/group). **(D)** HUDEP-2 cells were cultured in erythroid differentiation medium to differentiate. The mRNA levels of α, β and γ-globin in each group were quantified by Q-PCR. Untransduced cells (gray), SCR (blue), *MS4A3* shRNA (red). **(E)** Flow cytometric quantitation of mature red blood cells with Hoechst 33342 and GPA antibodies before and after HUDEP-2 cell differentiation (n=3/group). **(F)** Western blot analysis of CDK2 and p-CDK2 protein levels in untransduced cells, shSCR cells and *MS4A3* shRNA cells. β-actin was used as a loading control. Left, representative immunoblots; right, statistical analysis of gray value. The expression of CDK2 and p-CDK2 was normalized to that of β-actin to determine the ratios. **(G)** The mRNA levels of CDK2 in MEPs from the *Ad libitum* group and fasting group mice were measured by Q-PCR. *CDK2* mRNA levels were normalized to *Gapdh* (Feeding n=13; Fasting n=19). **(H)** The proportion of CDK2^+^ cells in MEPs from the *Ad libitum* group and fasting group mice was detected by multicolor flow cytometry (n=5/group). **(I)** p-CDK2 in MEPs was measured by flow cytometry in each group of mice (n=4/group). **(J)** Bone marrow MEP cells in each group of mice were measured using antibodies (Lin^−^Sca-1^−^c-kit^+^CD16/32^−^CD34^−^) on a polychromatic flow cell analyzer. (**K**) Flow cytometric quantitation of red blood cells with Ter119 antibody. (**L**) Peripheral blood cells by routine counting (Feeding + Vehicle n=10; Fasting + Vehicle n=10; Feeding + Roscovitine n=10; Fasting + Roscovitine n=9).

MS4A3 is a known negative regulator of the cell cycle upstream of cyclin-dependent kinase 2 (CDK2) that inhibits phosphorylation of the kinase (Donato, et al 2002; Chinami et al., 2005). In human HUDEPs, silencing *MS4A3* increased CDK2 expression and phosphorylation in MEPs, as measured by western blotting and flow cytometry (Figure 6F, S5A). *In vivo* mouse results indicated that downregulation of *Ms4a3* by fasting (Figure 5) is associated with both an increase in *Cdk2* transcription (Figure 6G) and CDK2-positive cells in mouse bone marrow MEPs (Figure 5H). To examine whether upregulation of *Cdk2* associated with fasting downregulation of *Ms4a3* expression is essential to promote the formation of RBCs, we used roscovitine, a CDK2 inhibitor, to treat the mice, and the results showed that rescovitine effectively inhibited p-CDK2-positive cells in bone marrow MEPs, and fasting was no longer able to increase p-CDK2-positive cells when mice were treated with roscovitine (Figure 6I). As expected, fasting-induced acceleration of the MEP cell cycle was blunted (Figure S5B). Accordingly, CDK2 inhibition blocked fasting-induced increases in the MEP pool (Figure 6J), and fasting failed to promote all types of bone marrow erythroid cells and ultimately the formation of mature red blood cells in the periphery when mice were treated *in vivo* with roscovitine (Figure 6K, L). The above data suggest that fasting inhibits the expression of *MS4A3* to activate CDK2 and thus accelerate MEP selfrenewal/proliferation and differentiation.

### 7. The response of erythropoietic MS4A3-CDK2 module to STIF depends on autophagy machinery, not EPO upregulation

Our recent proteomic analysis suggests *in vivo* autophagy activation by STIF in humans (Qian et al., 2021). *In vivo* autophagy in mice was also upregulated by 6 h fasting, as shown by increased expression of autophagy-essential genes (Figure 7A) and elevated markers for the formation of autophagosomes and autolysosomes (Figure 7B, S6A). The increase in erythropoiesis by STIF is comparable to that by autophagy induction with rapamycine (Figure 7C, S6B,C). To explore whether autophagy is implicated in STIF-enhanced erythropoiesis, we pharmacologically inhibited *in vivo* autophagy in mice with 3-methyladenine (3-MA), an inhibitor of phosphatidylinositol 3-kinases (Figure S6D,E). We found that fasting was no longer able to increase peripheral RBCs (Figure 7D, left panel) and bone marrow erythroid cells (Figure 7D, middle panel), and the fasting-enhanced proliferation of erythroblasts at stage IV was blocked (Figure 7D, right panel).

**Figure 7.**
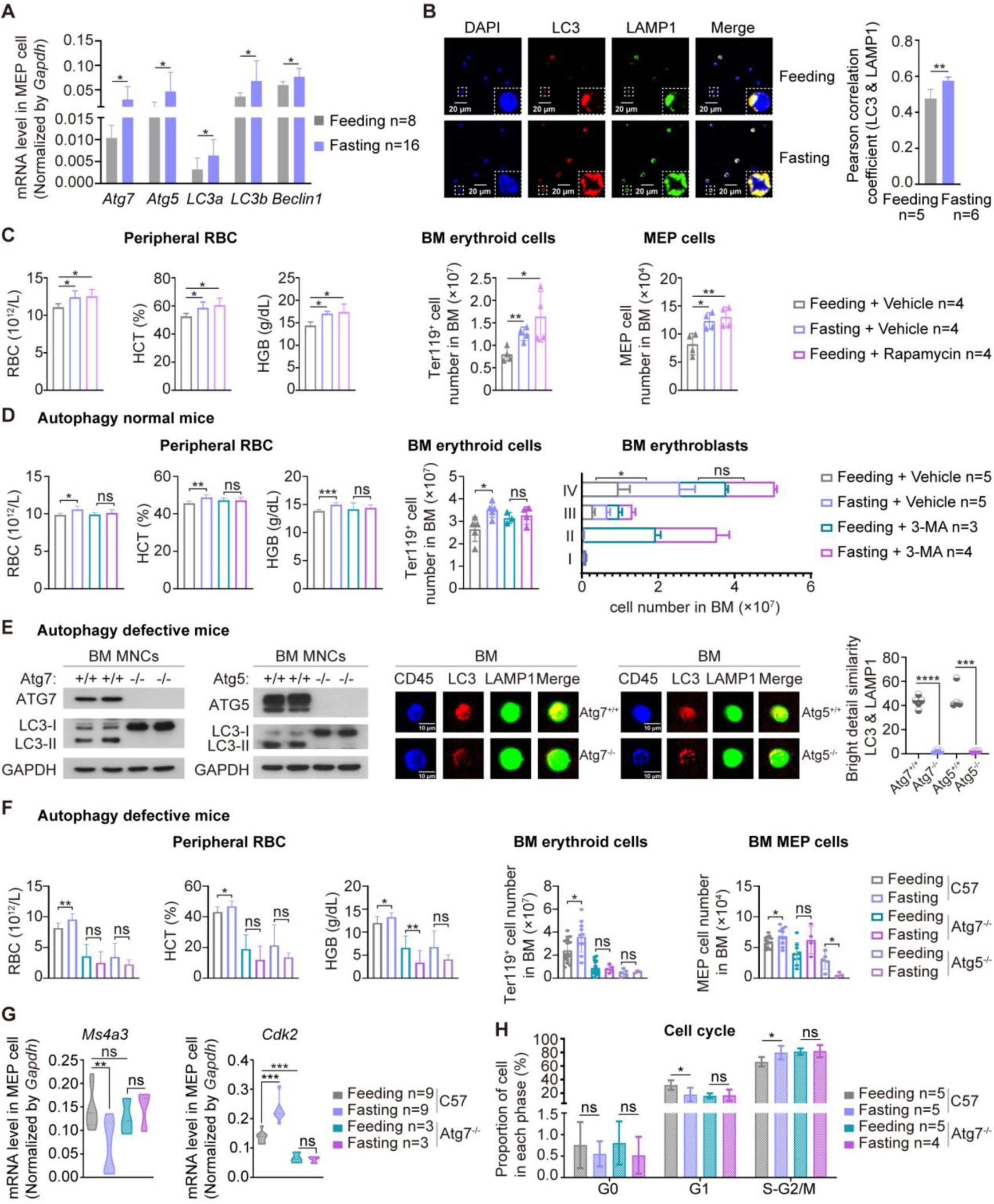
STIF-induced upregulation of erythropoiesis and downregulation of *Ms4a3* depend on intact autophagy machinery. **(A)** Expression of *Atg7*, *Atg5*, *LC3a*, *LC3b* and *beclin1* tested by Q-PCR. **(B)** Image flow cytometric analysis of the colocalization of DAPI, LC3 and LAMP1 was performed with an Amnis ImageStream Image Flow Cytometer. The Pearson correlation coefficient of LC3 and LAMP1 colocalization was calculated. (**C**) *In vivo* mouse treatment with rapamycin promoted erythropoiesis. Left, peripheral blood count from different treatments. Middle, Bone marrow erythrocyte count by flow cytometry. Right, MEP cells in bone marrow were measured on a polychromatic flow cell analyzer. (**D**) *In vivo* mouse treatment with 3-MA prevented fasting-promoted erythropoiesis. Left, peripheral blood count from different treatments. Middle, Bone marrow erythrocyte count by flow cytometry. Right, Flow cytometric determination of developmental stages in a bone marrow erythrocytic series using fluorescent antibodies against Ter119 and CD71. I: proerythroblast (Ter119^med^CD71^hi^); II: basophilic erythroblast (Ter119^hi^CD71^hi^); III: polychromatophilic erythroblast (Ter119^hi^CD71^med^); IV: orthochromatic erythroblast (Ter119^hi^CD71^−^). **(E)** Confirmation of autophagy defects in two autophagy-defective mouse models with *Atg7* or *Atg5* deleted in hematopoietic cells. Left, Western blotting of autophagy marker proteins. Right, Image flow cytometric analysis of the colocalization of autophagy marker proteins. **(F)** Hematopoietic cell counts in the different treatment autophagy-defective mouse groups. Left, peripheral blood cells by routine counting. C57 + Feeding (n=11); C57 + Fasting (n=10); *Atg7^−/−^* + Feeding (n=18); *Atg7^−/−^* + Fasting (n=8); *Atg5^−/−^* + Feeding (n=9); *Atg5^−/−^* + Fasting (n=3). Middle, bone marrow erythrocytes measured on polychromatic flow cell analyzer. C57 + Feeding (n=16); C57 + Fasting (n=11); *Atg7^−/−^*+ Feeding (n=16); *Atg7^−/−^* + Fasting (n=5); *Atg5^−/−^* + Feeding (n=5); *Atg5^−/−^* + Fasting (n=3). Right, Bone marrow MEP cells measured on polychromatic flow cell analyzer. C57 + Feeding (n=14); C57 + Fasting (n=11); *Atg7^−/−^* + Feeding (n=9); *Atg7^−/−^* + Fasting (n=4); *Atg5^−/−^* + Feeding (n=5); *Atg5^−/−^* + Fasting (n=3). **(G)** The expression of *Ms4a3* and *Cdk2* in MEP cells measured by quantitative PCR. **(H)** MEP cell cycle measured with Ki67-AF700 and Hoechst-33342 double fluorescent staining on a polychromatic flow cell analyzer.

To further confirm the importance of autophagy in STIF-enhanced erythropoiesis, we applied two mouse models with deletion of autophagy-essential gene *Atg*5 or *Atg*7 in hematopoietic system, tested by examination of *Atg* gene knockout by western blotting, LC3 lipidation and quantitative analysis of autophagosome formation by image flow cytometry (Figure 7E), and found that fasting was unable to enhance erythropoiesis since peripheral red blood cells, erythroid cells and MEP cells were unable to increase after fasting in either of the autophagy defective mice (Figure 7F). In particular, unlike wild-type MEPs, which were reduced in *Ms4a3* expression in response to 6 h fasting, autophagy-detective MEPs were unable to respond to downregulate *Ms4a3* or upregulate *Cdk2* (Figure 7G). In accordance with the above results, 6 h fasting was no longer able to accelerate the G1-S transition for a faster cell cycle in autophagy-defective mice (Figure 7H). These results suggest that the response of erythropoietic MS4A3-CDK2 module to STIF depends on autophagy.

Finally, to determine whether STIF-enhanced erythropoiesis requires upregulation of erythropoietin (EPO), we measured the expression of *Epo* and its receptor in mice before and after fasting. Fasting for 6 hours did not change the transcription level of *Epo* gene in the kidney or Epo protein level in the plasma of the fasting mice (Figure S7A,B), nor did 6 h fasting change the transcription level of Epo receptor gene (*EpoR*) in the mouse MEP cells (Figure S7C). In the human erythroid progenitor cell model HUDEP-2, increasing EPO concentration did not alter the expression levels of *MS4A3* or *CDK2*, nor did it alter the percentage of MS4A3 or p-CDK2 positive cells (Figure S7D,E). These data suggest that EPO upregulation is not required for STIF-enhanced erythropoiesis.

## DISCUSSION

In this study, we found that in humans, STIF, even occasionally practiced, can promote erythropoiesis and boost the function of RBCs in oxygen transportation, innate immunity, ATP production, and antioxidation. The impacts on the production and function of RBCs are relatively sustainable in the refeeding period following occasional STIF. Revitalization of erythropoiesis is more pronounced in adults with low RBC counts. Using mouse models and a human erythroid progenitor cell model, we proposed a mechanism by which STIF rejuvenates erythropoiesis. It involves selective regulation of MEPs by accelerating their selfrenewal and erythroid-biased differentiation without endangering normal hematopoiesis. Molecularly, STIF regulates an autophagy-dependent but EPO upregulation-independent MS4A3-CDK2 module to enhance erythropoiesis.

Aging humans show functional decline in hematopoiesis, and anemia is frequently diagnosed in older individuals (Ogawa, 2016; Jaiswal and Ebert, 2019; Jaiswal, 2020). Long-term dietary restriction and long-term modest fasting have been shown to decelerate the mammalian aging process (Longo et al., 2021) and result in positive therapeutic outcomes for certain diseases in both animal models and humans with or without combination chemotherapies (Lu et al., 2017; Cheng et al., 2017; Jordan et al., 2019; Caffa et al., 2020). Long-term caloric restriction delays HSC aging in mice (Chen et al., 2003) and T cell senescence in aged monkeys (Messaoudi et al., 2006). Prolonged fasting promotes HSC regeneration and reveres immunosuppression by reducing IGF-1/PKA in mice (Cheng et al., 2014).

Although long-term dietary restriction ameliorated HSC aging phenotypes, such as the increase in the number of HSCs and the skewing toward myeloid-biased HSCs during aging and increased HSC quiescence and improved the maintenance of the repopulation capacity of HSCs during aging, long-term dietary restriction strongly impaired HSC differentiation into lymphoid lineages and particularly inhibited the proliferation of lymphoid progenitors, resulting in decreased production of peripheral B lymphocytes and impaired immune function in mice (Poole et al., 2016; Tang et al., 2016), suggesting a potential risk for long-term feeding intervention. Nevertheless, positive responses from caloric restriction mimetics triggering an anticancer immune response in mice (Pietrocola et al., 2016) and from fasting-mimicking diet combined with chemotherapy enhancing killing of human breast cancer cells (Di Biase et al., 2016) and from intermittent fasting improving chemotherapy on acute lymphoblastic leukemia in mice (Lu et al., 2017), as well as from intermittent fasting blunting human CD4^+^ T helper cell responsiveness to reduce inflammatory level (Han et al., 2021) encourage intervention in human subjects by STIF, which has been much less explored in hematopoiesis.

Our recent study found that for low platelet count subjects, STIF brought about a quite sustainable elevated average platelet count for better hemostasis, whereas for subjects with normal platelet counts, STIF practice led to a progressive decrease in its count to a lower value within the normal range to reduce thrombosis risk without compromising hemostasis capacity, reflecting the human body’s adaptive and protective response to intensive fasting (Fang et al., 2021). We also observed that STIF leads to innate immune remodeling, particularly with improved function of neutrophils (Qian et al., 2021).

Here, we show that STIF not only promotes the production of red blood cells, but also improves the function of red blood cells in both oxygen transportation and innate immune response, which is in support of a recent finding showing immunomodulatory capacity of human red blood cells by single-cell transcriptomic analysis (Xu et al., 2022).

Erythropoiesis is regulated at multiple levels to ensure effective production of RBCs (Caulier & Sankaran, 2021). MS4A3 has been reported to be a hematopoietic cell cycle modulator. Specifically, it regulates the G1-S cell cycle transition by phosphorylating CDK2 or through its direct binding to CDK inhibitor 3 in the nucleus (Donato et al. 2002; Chinami et al., 2005; Kutok et al., 2005). In our study, however, we found that silencing *MS4A3* in human erythroid progenitor HUDEP cells led to upregulation of both *CDK2* gene transcription and CDK2 protein phosphorylation. In mice, downregulation of *Ms4a3* transcription by STIF is associated with increased transcription of the *Cdk2* gene and phosphorylation of the Cdk2 protein. These results indicate that MS4A3 regulates CDK2 not only at the posttranslational modification level but also at the transcription level, suggesting a unique modulatory interplay between MS4A3 and CDK2 in a STIF context.

Autophagy is an evolutionarily conserved mechanism that digests unnecessary or dysfunctional cellular components and allows survival by intercellular self-eating cell metabolism (Galluzzi et al., 2014). In model animals, autophagy was found to be essential for HSC maintenance and multilineage differentiation (Mortensen et al., 2011; Cao et al., 2015a; 2015b) and to improve the lifespan-prolonging effects of calorie-limited diets (Fernandez et al., 2018; Ulgherait et al., 2021). A periodic fasting-mimicking diet prevented the age-dependent accumulation of p62, a marker of reduced autophagy activity, which suggests that the healthspan-promoting effects of such fasting are carried out at least in part by activation of autophagy (Brandhorst et al., 2015). In mice, basal autophagy is found to clear excessive or old mitochondria during reticulocyte maturation (Schweers et al., 2007; Sandoval et al., 2008; Kundu et al., 2008; Zhang et al., 2009; Mortensen et al., 2010; Li-Harms et al., 2015). Thus, the dysfunction of autophagy hinders correct erythroid maturation, leading to anemia in mice. However, the molecular link between autophagy and erythroid maturation remains unidentified in both mice and humans.

Our recent studies found that autophagy in mice and humans collaborates with Sirt3, an anti-aging protein, to decelerate hematopoietic aging (Fang et al., 2020) and that STIF induces *in vivo* autophagy in human subjects (Qian et al., 2021). Here, using human erythroid progenitor cell and mouse models, we found that STIF-induced downregulation of MS4A3 and upregulation of CDK2 that together confer erythropoiesis all depend on autophagy integrity. Without fasting, *Ms4a3* did not show a difference in basal transcription between wild-type and *Atg7*-deleted MEPs, but *Ms4a3* downregulation was highly dependent on the autophagy machinery since fasting was no longer able to reduce *Ms4a3* expression in mice with autophagy defects in the hematopoietic system. In contrast, *Cdk2* upregulation highly relies on autophagy machinery since the *Cdk2* transcript of *Atg7*-deleted MEPs is much lower than that of wild-type mice, and autophagy defects disable STIF-triggered upregulation of *Cdk2* expression and Cdk2 phosphorylation.

A long-established dogma is that upregulation of erythropoietin (EPO) is responsible for increased erythropoiesis, and hypoxia is a strong trigger for EPO expression (Eckardt et al., 1989; Schuster et al., 1989; Ge et al., 2002; Robach et al., 2004; Watts et al., 2020). However, in the present study we did not find hypoxia during STIF practice. A recent study shows that without fasting stress, mitochondrial autophagy regulates Epo production and protects against renal anemia (Geng et al., 2021), also suggesting a dependency on EPO upregulation for erythropoiesis in steady physiological condition. In contrast, our results demonstrate that the STIF-induced MS4A3-CDK2 module regulates erythropoiesis independent of EPO upregulation, suggesting an unusual mechanism underlying the response of erythropoiesis to STIF intervention.

Therefore, our study indicates that STIF, as a nonmedical intervention, triggers rejuvenation of erythropoiesis with a unique mechanism independent of EPO upregulation but dependent on the autophagy-supported MS4A3-CDK2 module. Most importantly, the combined results from STIF interventions in multiple cities in China indicate that subjects with low RBC counts can achieve more significant rejuvenation of erythropoiesis, suggesting that occasional STIF can be more effectively practiced in such subhealthy populations.

## STAR METHODS

- KEY RESOURCES TABLE
- RESOURCE AVAILABILITY

- Lead contact
- Material availability
- Data availability
- EXPERIMENTAL MODEL and SUBJECT DETAILS

- Mouse Models
- Cell culture
- METHODS DETAILS

- Study design
- Blood routine analysis
- Morphological detection of RBCs
- Assays for ATP, SOD, CAT, MDA, GSH-Px, 2,3-DPG and Pfhb
- Flow cytometry and cell sorting
- Cell proliferation analysis
- Apoptosis analysis
- Cell cycle analysis in MEP cells
- CFU assay
- Quantitative Real-time PCR analysis
- Western blot analysis
- Lentivirus-mediated RNA Interference
- ImageStream analysis
- Immunofluorescence
- RNA sequencing in mouse MEP cells
- Proteome sequencing in human RBCs
- QUANTIFICATION AND STATISTICAL ANALYSIS

- Data representation and statistical analysis

## Data Availability

All data produced in the present work are contained in the manuscript

## ACKNOWLEDGMENTS

We thank all the volunteers participating in this fasting study and the campus Hospital of Soochow University for providing the facilities in the collection of blood samples and taking physical measurement. This study was supported in part by the National Natural Science Foundation of China by grants No.91649113, No.82170227 and No.31771640 (to JW), No.82000117 (to YF), No.81800152 (to NY), No.81673093 (SZ) and by Soyo Center of Soochow University (to JW) by grant H190443, and by the Natural Science Foundation of Jiangsu Province, China (to YF) by grant BK20200191, and by the Postdoctor Science Foundation of Jiangsu Province (to YF) by grant 2020Z064, and the Priority Academic Program Development of Jiangsu Higher Education Institutions, the Project of State Key Laboratory of Radiation Medicine and Protection, Soochow University by grant No. GZC00201(to JW), the Aerospace Medical Experiment Project of Space Station by grant No.HYZHXM02002.

## AUTHOR CONTRIBUTIONS

J.W. conceived and supervised the study. J.W., Y.F., N.Y., S.Z., L.X., W.W., Y.L., L.W., L.L., Y.G., C.Z. and X.G. organized human fasting interventions and collected blood samples. L.X., N.Y., J.Q. and Y.F designed the experiments. L.X. performed the most of the experiments. Z.J., X.W., W.W., Y.G., W.B., Y.Y., R.Z. and X.M participated in the experiments. Z.Y., Z.C., Y.X., P.X., Y.Z. provided the reagents. L.X. analyzed most of the data; J.Q. performed transciptomic and proteomic analysis. L.X. and J.Q. drafted the manuscript; J.W., Y.N. and Y.F. edited the manuscript.

## INCLUSION AND DIVERSITY

We worked to ensure sex balance in the enrollment of human fasting subjects and in the selection of no-human fasting subjects.

## STAR METHODS

### KEY RESOURCES TABLE

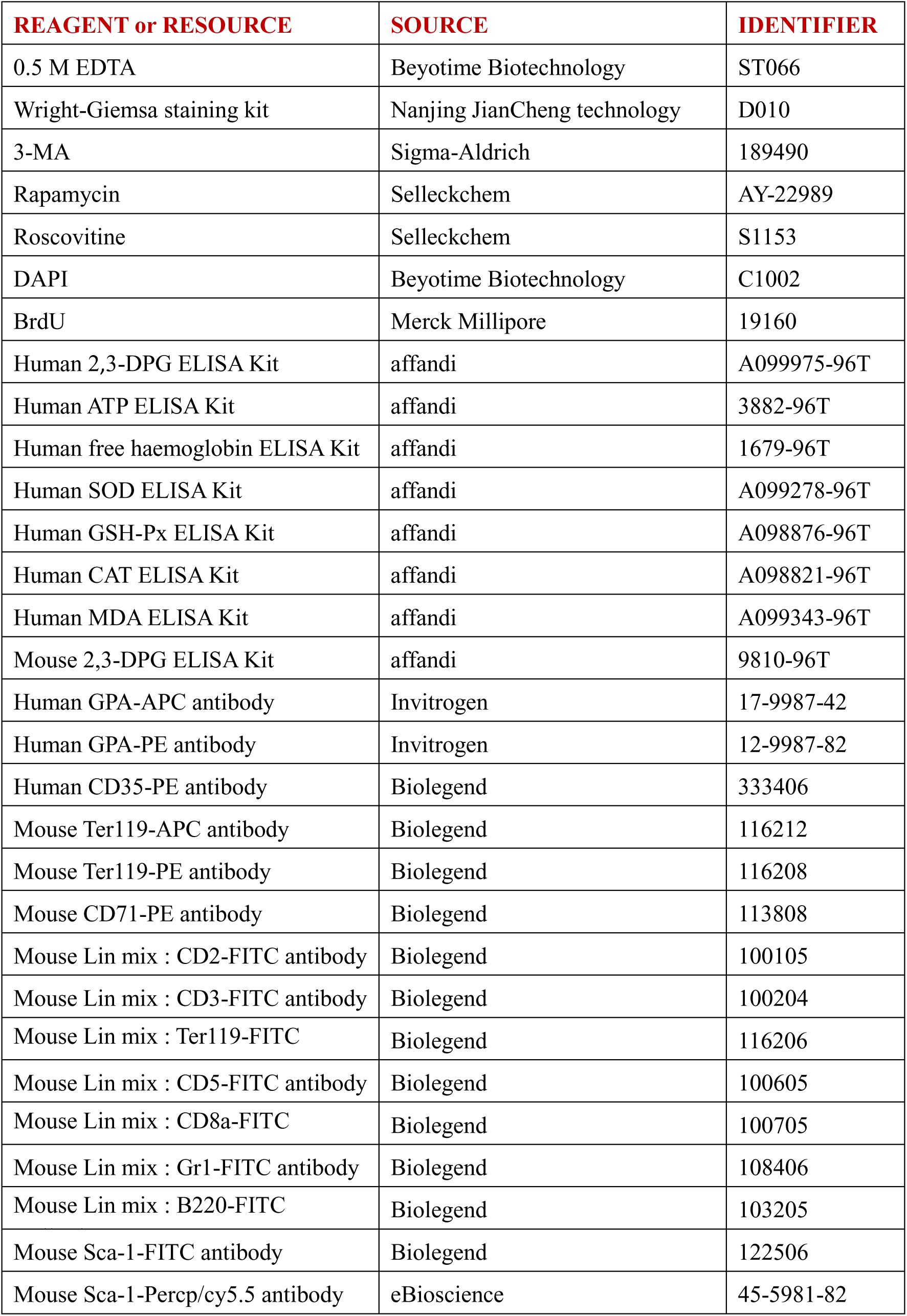
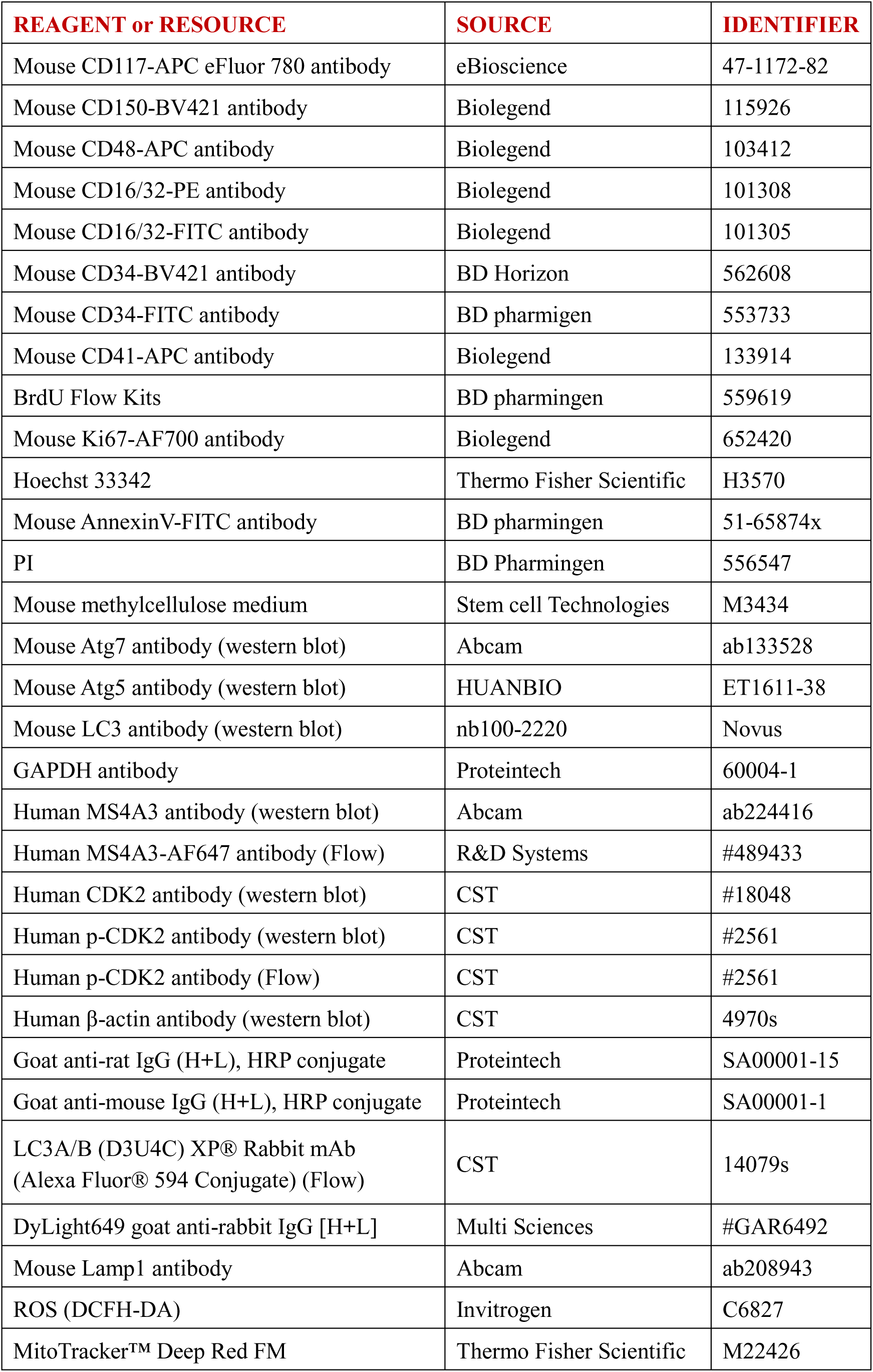
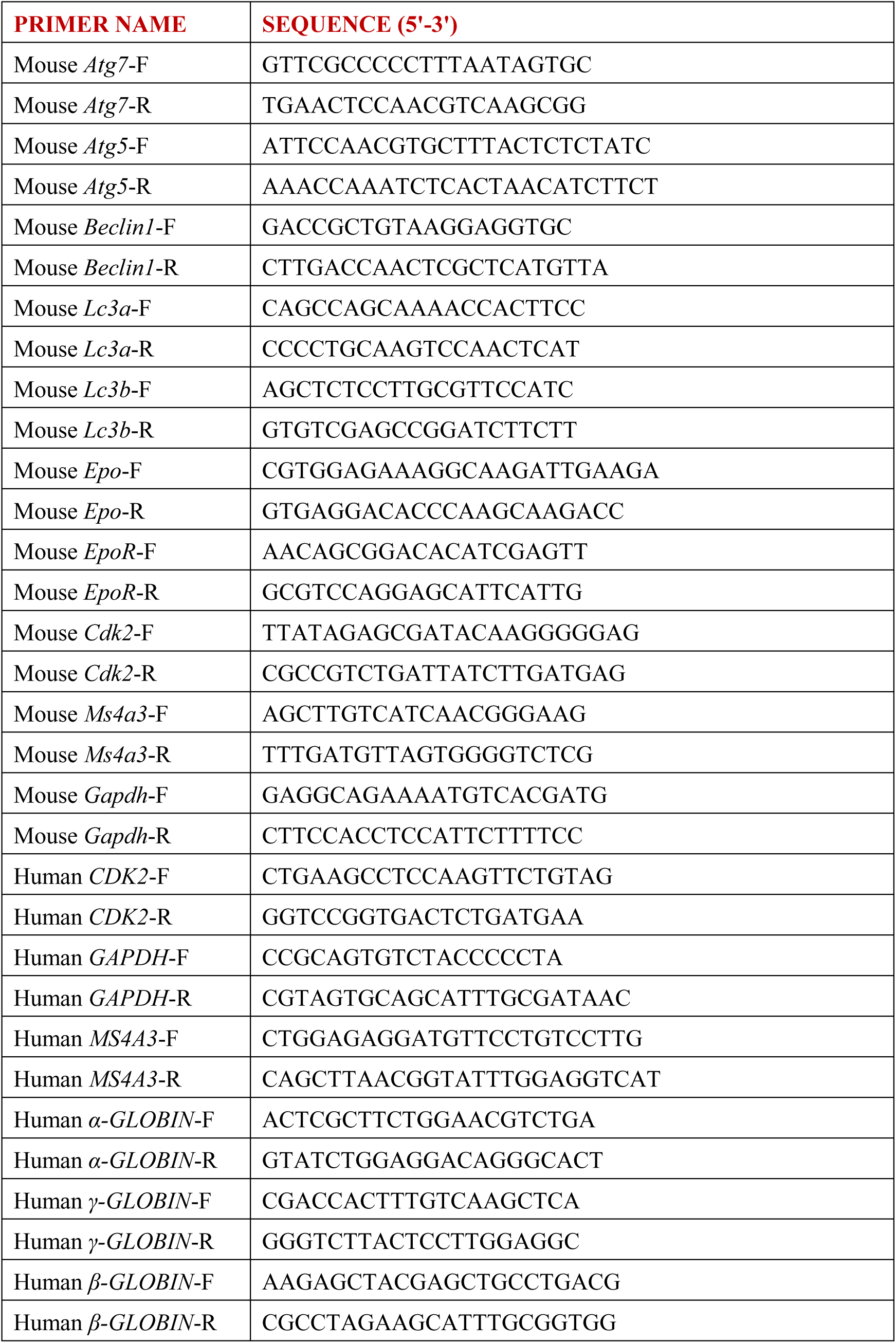

### RESOURCES AVAILABILITY

#### Lead contact

Further information and requests for resources and reagents should be directed to and will be fulfilled by the lead contact, Jianrong Wang.

#### Materials availability

Mouse models used in this study are commercially available.

#### Data and code availability

- Raw RNA sequencing data for mouse MEPs have been deposited at Gene Expression Omnibus under the accession number GSE200918. Raw proteomics data for human red blood cells is available upon request.
- riginal western blot images, flow cytometry data, and microscopy data reported in this paper will be shared by the lead contact upon request.
- This paper does not report original code.
- Any additional information required to analyze the data reported in this paper is available from the lead contact upon request.

### EXPERIMENTAL MODEL AND SUBJECT DETAILS

#### Study design

The study was conducted with approval from the Institutional Ethics Review Board at Soochow University (Approval No. ECSU-2019000153) and the Chinese Clinical Trial Register, an official review board for clinical trials (Registration No. ChiCTR1900027451), affiliated with The Ministry of Public Health of China (http://www.chictr.org.cn/index.aspx). Participants were enrolled after giving their written informed consent. Thirty-one participants aged 27 to 67 years were enrolled. Exclusion criteria and fasting programs as well as refeeding protocols were described previously (Fang et al., 2021). Blood samples were collected at 7:00 to 9:00 am on testing days indicated in the figures.

#### Mice

C57BL/6 mice and Vav-iCre mice were purchased from the Jackson Laboratory. The generation of Atg7^floxp/floxp^ mice has been previously described (Komatsu et al., 2005; Cao et al., 2015; Fang et al., 2021). Atg5^floxp/floxp^ mice were provided by Dr. Noboru Mizushima, Japan (Mizushima et al., 2004). All animal experiments were reviewed and approved by the Institutional Committee on Animal Welfare Protection and Ethics of Soochow University.

#### Chemicals and biological reagents

3-MA (189490) was purchased from Sigma-Aldrich. To inhibit autophagy, C57BL/6 mice were intraperitoneally injected with 15 mg/kg 3-MA for two days. Rapamycin (AY-22989) was purchased from Selleckchem. To activate autophagy, C57BL/6 mice were intraperitoneally injected with 2 mg/kg rapamycin for one week. Roscovitine (S1153), a p-CDK2 inhibitor, was purchased from Selleckchem. 100 mg/kg roscovitine was intraperitoneally injected to C57BL/6 mice for two days. Detailed information on the biological reagents is provided in Key RESOURCES TABLE in the Supplementary Information.

#### Blood routine analysis

First, we prepared the blood thinner. The blood thinner included 100 μL EDTA (ST066, Beyotime Biotechnology) and 50 μL PBS. Then, 150 μL peripheral blood (PB) of each group of mice was collected. Finally, we performed routine five-class blood analysis and analyzed the RBC counts, hemoglobin concentration and hematocrit of each group of mice.

#### Morphological detection of RBCs

RBCs were obtained from PB by centrifugation at 3000 rpm/min for 10 min. RBCs were transferred to glass slides by centrifugation at 400 rpm/min for 5 min. A Wright-Giemsa stain kit (D010, Nanjing JianCheng Technology) was used to detect the morphology of the RBCs according to the manufacturer’s instructions.

#### ATP, SOD, CAT, MDA, GSH-Px, 2,3-DPG

We collected the PB of each group mice. RBCs were obtained by centrifugation at 3000 rpm/min for 10 min. The concentrations of ATP, SOD, CAT, MDA, GSH-Px and 2,3-DPG were determined using a human ELISA Kit (affandi) on a Labsystem Multiskan MS (Thermo) according to the manufacturer’s instructions.

#### Pfhb

Pfhb levels in plasma were assayed by enzyme-linked immunosorbent assay (ELISA) using a human Pfhb ELISA Kit (affandi) according to the manufacturer’s instructions.

#### Flow cytometry and cell sorting

Single-cell suspensions from BM and PB were stained with panels of fluorochrome-conjugated antibodies (Table S1). RBCs from PB in humans were stained with GPA-PE (12-9987-82, Invitrogen) antibody. RBCs from BM and PB in mice were stained with Ter119-APC (116212, Biolegend) and CD71-PE (113808, Biolegend) antibodies. HSCs were labeled with lineage FITC (Biolegend), Sca-1-Percp/Cy5.5 (45-5981-82, eBioscience), CD117-APC-eFluor 780 (47-1172-82, eBioscience), CD150-BV421 (115926, Biolegend) and CD48-APC (103412, Biolegend) antibodies. MEP cells were labeled with lineage-Fitc, Sca1-Percp/Cy5.5, CD117-APC-eFluor 780 (47-1172-82, eBioscience), CD16/32-PE (101308, Biolenged) and CD34-BV421 (562608, BD Horizon) antibodies. The analyses were performed using Beckman Coulter gallios. All data were analyzed using Kaluza. Information on the antibodies is provided in Key RESOURCES TABLE.

#### Cell proliferation analysis

BrdU (100 mg/kg, 19160, Merck Millipore) was injected daily into the peritoneum for 7 days, with the last injection 6 hours prior to harvesting BM. BrdU-positive nuclei were detected with BD Pharmingen™ BrdU Flow Kits (559619, BD Pharmingen) according to the manufacturer’s instructions. Ki67-AF700 (652420, Biolegend)-labeled cells were counted in MEP cells by flow cytometry.

#### Apoptosis analysis

RBCs in PB were labeled with Ter119-PE (116208, Biolegend) and Annexin V-FITC (51-65874x, BD Pharmingen) antibodies. RBCs in BM were labeled with Ter119-BV510 (116237, Biolegend), Annexin V-FITC and PI (556547, BD Pharmingen). Apoptotic cells (Annexin V^+^ PI^−^) were detected by flow cytometry (Beckman Coulter gallios).

#### Cell cycle analysis in MEP cells

MEP cells were labeled with lineage FITC (Biolegend), Sca-1 FITC (108106, Biolegend), c-kit APC-eFluor 780 (47-1172-82, eBioscience), CD34-FITC (553733, BD Pharmingen) and CD16/32-FITC (101305, Biolegend) antibodies. Ki67-AF700 (652420, Biolegend) and Hoechst-33342 (H3570, Thermo Fisher Scientific) were used to detect the cell cycle in MEP cells.

#### CFU assay

BM cells were collected from the mice in the *ad libitum* group and fasting group. A total of 10000 cells were plated in drug-free methylcellulose medium (M3434, Stem Cell Technologies). Colonies were counted on day 7.

#### Quantitative Real-time PCR analysis

Total RNA was extracted with a MicroElute Total RNA Kit (R6831, Omega), and cDNA was synthesized using a revertaid cDNA synthesis kit (K1622, Thermo Fisher Scientific) according to the manufacturer’s instructions. RT-qPCR was performed in triplicate using a Roche LightCycler 480 Instrument II with LightCycler 480 SYBR Green I Master Mix (04707516001, Roche). Expression of the gene of interest was normalized to the housekeeping gene GAPDH. All RT-qPCR primers used are listed in Key RESOURCES TABLE in the Supplementary Information.

#### Western blot analysis

Cells were harvested from 8-week-old C57BL/6 mice. Cells were lysed in lysis buffer (Cell Signaling) containing protease inhibitor mixture and PhosSTOP phosphatase inhibitor mixture on ice for 30 min. Crude lysates were obtained by centrifugation centrifuged at 13,000 g for 20 min and heated at 95°C for 5 min at 4°C. The protein was detected by immunoblotting. Equal amounts of protein were loaded on a 12% SDS-polyacrylamide gel electrophoresis (SDS-PAGE) and transferred to polyvinylidene difluoride membranes (Millipore). ATG7 (ab133528, Abcam), ATG5 (ET1611-38, HUABIO), LC3 (nb100-2220, Novus), GAPDH (60004-1, Proteintech), MS4A3 (ab224416, Abcam), CDK2 (#18048, CST), p-CDK2 (#2561, CST) and β-actin (4970s, CST) antibodies were revealed using an appropriate horseradish peroxidase (HRP)-conjugated secondary antibody (Cell Signaling) and detected by an enhanced chemiluminescence kit (Pierce). Secondary antibodies were purchased from Cell Signaling Technology. Blots were probed with anti-GAPDH antibody to confirm equal loading of protein.

#### Lentivirus-mediated RNA Interference

Lentiviral vectors to silence MS4A3 together with negative control vector (shRNA-NC) were purchased from GenePharma Co, Ltd. (Shanghai, China). The sequences to silence *MS4A3* were shRNA-1(5-GGTTCCTTGCAATACCCATAC-3), shRNA-2(5-GGCCTCCCACGAAGTTGATAA-3), and shRNA-3(5-GGGACCAGAAGAGCTGAATAC-3). Virus was produced by cotransfection of 293T cells with sh*MS4A3* or shRNA-NC and packaging plasmids. For viral infection, viral supernatant was filtered, collected and used to infect HUDEP-2 cells. HUDEP-2 cells were cultured in Stemspan SFEM medium (#09655, Stem Cell Technologies), supplemented with 1% penicillin/streptomycin (AP231, AS325, Beijing Dingguo Changsheng Biotechnology), 50 ng/mL SCF (#300-07, Peprotech), 3 IU/mL EPO (#100-64, Peprotech), 1 μM dexamethasone (#D4902, Sigma-Aldrich) and 1 μg/mL doxycycline (#4090/50, R&D Systems) for 48 h. Lentivirus was then added to the culture medium. After 72 h, GFP^+^ HUDEP-2 cells were sorted for western blot and flow cytometry.

#### ImageStream analysis

Image flow cytometric analysis of colocalization of CD45-FITC (157214, Biolegend), LC3A/B Alexa Fluor® 594 Conjugate (14079s, CST) and Lamp1 (ab208943, Abcam) was performed with an Amnis ImageStream Image Flow Cytometer (Amnis, Merck Millipore). First, cells were stained with CD45-FITC (diluted 1:100) for 30 min. After fixation and permeabilization using 4% PFA and 0.1% saponin (S7900, Sigma), cells were stained with primary antibodies against LC3 and LAMP1 (diluted 1:100) for 30 min and then stained with DyLight 649 goat anti-rabbit IgG [H+L] for 30 min. Finally, cells were explored on an ImageStreamX Mark II machine for image flow cytometry. Samples were visualized and analyzed for the expression of marker proteins with IDEAS 6.0 software (Amnis, Merck Millipore). Scale bar: 10 μm.

#### Immunofluorescence

MEP cells were first sorted by flow cytometry and were then fixed in 4% paraformaldehyde for 15 minutes, permeabilized in 0.5% Triton X-100 (DH351-2, Beijing Dingguo Changsheng Biotechnology) in PBS for 5 minutes, blocked with 4% goat serum in PBS for 60 minutes, and incubated with LC3A/B Alexa Fluor® 594 Conjugate and Lamp1 overnight at 4°C. A DyLight 649 goat anti-rabbit IgG [H+L] antibody was used as a secondary antibody. Samples were stained with DAPI (C1002, Beyotime) before photographing under a fluorescence microscope (FV1000MPE-share). The colocalization of LC3 and LAMP1 in MEP cells was analyzed by Cell sense.

#### RNA sequencing in mouse MEP cells

##### 1) Total RNA extraction

Total RNA was extracted from the tissues using Trizol (Invitrogen, Carlsbad, CA, USA) according to the manufacturer’s instructions. Approximately 60 mg of tissue was ground into powder in liquid nitrogen in a 2 mL tube, homogenized for 2 minutes and rested horizontally for 5 minutes. The mix was centrifuged for 5 minutes at 12,000×g at 4°C, and then the supernatant was transferred into a new EP tube with 0.3 mL chloroform/isoamyl alcohol (24:1). The mix was shaken vigorously for 15 s and then centrifuged at 12,000×g for 10 minutes at 4°C. After centrifugation, the upper aqueous phase where RNA remained was transferred into a new tube with an equal volume of isopropyl alcohol supernatant and then centrifuged at 13,600 rpm for 20 minutes at 4°C. After deserting the supernatant, the RNA pellet was washed twice with 1 mL 75% ethanol, and then the mix was centrifuged at 13,600 rpm for 3 minutes at 4°C to collect residual ethanol, followed by pellet air drying for 5-10 minutes in a biosafety cabinet. Finally, 25 µL∼100 µL of DEPC-treated water was added to dissolve the RNA. Subsequently, total RNA was qualified and quantified using a NanoDrop and Agilent 2100 bioanalyzer (Thermo Fisher Scientific, MA, USA).

##### 2) mRNA Library Construction

Oligo(dT)-attached magnetic beads were used to purify mRNA. Purified mRNA was fragmented into small pieces with fragment buffer at the appropriate temperature. Then, first-strand cDNA was generated using random hexamer-primed reverse transcription, followed by second-strand cDNA synthesis. Afterwards, A-Tailing Mix and RNA Index Adapters were added by incubation for end repair. The cDNA fragments obtained from the previous step were amplified by PCR, and the products were purified by Ampure XP Beads and then dissolved in EB solution. The product was validated on an Agilent Technologies 2100 bioanalyzer for quality control. The double-stranded PCR products from the previous step were heated, denatured and circularized by the splint oligo sequence to obtain the final library. Single strand circle DNA (ssCir DNA) was formatted as the final library. The final library was amplified with phi29 to make DNA nanoballs (DNBs), which had more than 300 copies of one molecule. DNBs were loaded into the patterned nanoarray, and paired-end 100-base reads were generated on the BGISeq500 platform (BGI-Shenzhen, China).

##### 3) Reference genome and annotation

The human genome sequence (primary assembly, GRCm38.p6) fasta file, comprehensive gene annotation GTF file (primary assembly, GENCODE release M25), and metadata (UniProtKB/SwissProt entry associated with the transcript, GENCODE release M25) were downloaded from the GENCODE website. The index for HISAT2 was built using GENCODE genome sequences according to the hisat2-build manual. Gene features (gene length, gene-transcript-swiss prot-id conversion) were generated from the GTF file using in-house scripts.

##### 4) RNA seq data analysis

All mouse MEP-sequenced fastq files were generated and cleaned by BGI, and then the RNA sequencing reads were aligned using HISAT2 (version 2.2.1) (Kim et al., 2019) with a self-built ht2 index. The parameters were default for paired-end analysis. Aligned reads were stored in SAM files. Next, SAM files were sorted into BAM files as well as for compression using samtools (version 1.11) (Danecek et al., 2021). For quantification, feature Counts (Liao et al., 2013) from the Subread package (version 2.0.1) was used with comprehensive gene annotation GTF with exon as the feature type and with gene id as the meta feature. Differentially expressed gene analysis was performed using the R package DESeq2 (version 1.30.0) (Love et al., 2014). We determined DEGs with |log_2_FoldChange| > 1 and adjusted *P* value < 0.05.

#### Proteome Sequencing in human RBCs

##### 1) Protein extraction and quality control

Take an appropriate amount of sample, transfer it to a 1.5 mL centrifuge tube, add an appropriate amount of protein lysis buffer containing SDS (sodium dodecyl sulfate) and a final concentration of 1x protease inhibitor cocktail with EDTA; ultrasound in ice bath, centrifuge at 25, 000 g 4°C for 15 min, take the supernatant and add DTT (dithiothreitol) with a final concentration of 10 mM, and place in a water bath at 37°C for 30 min; add IAM (Iodoacetamide) with a final concentration of 55 mM, and place in a dark room for 45 min; add 5 times the volume of precooled acetone, place in −20°C refrigerator for 2 h, centrifuge at 25, 000 g 4°C for 15 min and discard the supernatant; air-dry the precipitation, add an appropriate amount of SDS-free protein lysis solution, and use an automatic grinder to promote protein dissolution; centrifuge at 25, 000 g 4°C for 15 min and take the supernatant, which is the protein solution. Standard proteins (0.2 μg/μL BSA) 0, 2, 4, 6, 8, 10, 12, 14, 16, 18 μL were sequentially added to the 96-well microtiter plates A1 to A10, followed by the addition of pure water 20, 18, 16, 14, 12, 10, 8, 6, 4, 2 μL, and then 180 μL of Coomassie Brilliant Blue G-250 Quantitative Working Solution was added to each well. The OD595 was measured with a microplate reader, and a linear standard curve was drawn based on the OD595 and protein concentration. The protein solution was diluted to be tested several times, 180 μL of the quantitative working solution was added to 20 μL of the protein solution, and the OD595 was read. The sample protein concentration was calculated from the standard curve and sample OD595. Each 10 μg of protein solution was mixed with an appropriate amount of loading buffer, heated at 95°C for 5 minutes, and centrifuged at 25,000 g for 5 minutes, and the supernatant was loaded into a well of a 12% SDS polyacrylamide gel. After electrophoresis, Coomassie blue staining was carried out for 2 hours, after which an appropriate amount of decolorizing solution (40% ethanol 10% acetic acid) was added to the shaker to decolorize 3 to 5 times for 30 minutes each time. One hundred micrograms of protein solution per sample was diluted with 50 mM NH_4_HCO_3_ in 4 volumes. Two micrograms of trypsin enzyme was added at a protein:enzyme ratio of 40:1 and digested for 4 hours at 37°C. Enzymatic peptides were desalted using a Strata X column and vacuumed to dryness.

##### 2) High pH RP separation

Equal amounts of peptides were extracted from all samples to mix, and the mixture was diluted with mobile phase A (5% ACN pH 9.8) and injected. A Shimadzu LC-20AB HPLC system coupled with a Gemini high pH C18 column (5 μm, 4.6 x 250 mm) was used. The sample was subjected to the column and then eluted at a flow rate of 1 mL/min by gradient: 5% mobile phase B (95% ACN, pH 9.8) for 10 minutes, 5% to 35% mobile phase B for 40 minutes, 35% to 95% mobile phase B for 1 minute, flow Phase B lasted 3 minutes and 5% mobile phase B equilibrated for 10 minutes. The elution peak was monitored at a wavelength of 214 nm, and the components were collected every minute. Components were combined into a total of 10 fractions, which were then freeze-dried.

##### 3) DIA quantification (Nano-LC–MS/MS)

The dried peptide samples were reconstituted with mobile phase A (2% ACN, 0.1% FA), centrifuge at 20,000 g for 10 minutes, and the supernatant was taken for injection. Separation was carried out by a Thermo UltiMate 3000 UHPLC liquid chromatograph. The sample was first enriched in the trap column and desalted and then entered a tandem self-packed C18 column (150 μm internal diameter, 1.8 μm column size, 35 cm column length) and separated at a flow rate of 500 nL/min by the following effective gradient: 0∼5 minutes, 5% mobile phase B (98% ACN, 0.1% FA); 5∼90 minutes, mobile phase B linearly increased from 5% to 25%; 90∼100 minutes, mobile phase B rose from 25% to 35%; 100∼108 minutes, mobile phase B rose from 35% to 80%; 108∼113 minutes, 80% mobile phase B; 113.5∼120 minutes, 5% mobile phase B. The nanoliter liquid phase separation end was directly connected to the mass spectrometer as the following settings. The peptides separated by liquid phase chromatography were ionized by a nanoESI source and then passed to a tandem mass spectrometer Oritrap Exploris 480 (Thermo Fisher Scientific, San Jose, CA) for DDA (Data were ionized Acquisition) detection mode. The main parameters were set: ion source voltage was set to 1.9 kV, MS1 mass spectrometer scanning range was 400∼1,250 m/z; resolution was set to 120,000; maximal injection time (MIT) 90 ms; 400∼1,250 m/z was equally divided to 50 continuous windows MS/MS scan. MS/MS collision type HCD, collision energy NCE 30; MIT was auto mode. Fragment ions were scanned in Orbitrap at an MS/MS resolution of 30,000. AGC was: MS 300%, MS/MS 1000%. The DIA data were analyzed using the iRT peptides for retention time calibration. Then, based on the target-decoy model applicable to SWATH-MS, false positive control was performed with FDR 1%, therefore obtaining significant quantitative results.

##### 4) MSstats differential analysis

MSstats is an R package from the Bioconductor repository. It can be used for statistical evaluation of significant differences in proteins or peptides from different samples and is widely used in targeted proteomics MRM, label-free quantitation, and SWATH quantitative experiments. The core algorithm is linear mixed effect model. The process preprocessed the data according to the predefined comparison group and then performed the significance test based on the model. Thereafter, differential protein screening was performed based on a fold change >1.5 and a *P* value<0.05 as the criteria for significant differences. At the same time, enrichment analysis was performed on the differentially expressed proteins.

##### 5) Gene ontology enrichment analysis

The Gene Ontology OBO file (release 2021-02-01) was downloaded from geneontology.org. The human Gene Ontology annotation GAF file (release 2021-02-01) was downloaded from the EBI Gene Ontology Annotation Database. We conducted overrepresentation analysis on DEGs/DEPs identified from differential analysis using the python library GOATOOLS. All DEG enrichment query lists were converted from Ensemble gene id to protein id. Enrichment background was generated from the gene ontology annotation GAF file using an in-house script. The criterion for identifying significantly overrepresented terms was set as an adjusted P value < 0.05.

##### 6) Gene set enrichment analysis

We conducted gene set enrichment analysis (GSEA) on preranked gene lists with log2FoldChange as the ranking metric using GSEA v4.1.0 (Subramanian et al., 2005). We used gene ontology terms as gene sets, and the gene sets were generated using an in-house script from gene ontology analysis.

### QUANTIFICATION AND STATISTICAL ANALYSIS

#### Data representation and statistical analysis

For human data, **s**tatistical analyses were performed using SPSS version 22.0. Differences between groups were analyzed using univariate repeated-measures analysis of variance (RM ANOVA). To assess changes during fasting, we performed each fasting time point to baseline (before fasting) using one-way analysis of variance (ANOVA) with multiple comparisons. Multiple comparisons were performed using Dunnett’s test and then using the Benjamini & Hochberg method to control the false discovery rate. Data were expressed as the mean ± standard deviation (SD). Adjusted *P*<0.05 was considered to indicate a statistically significant difference.

For mouse data, **s**tatistical analyses were performed using Graphpad Prism 8. The statistical significance of the observed differences was determined by unpaired Student’s t test. Data were expressed as the mean ± standard deviation (SD). *P*<0.05 was considered to indicate a statistically significant difference.

## SUPPLEMENTARY INFORMATION

**Figure S1.**
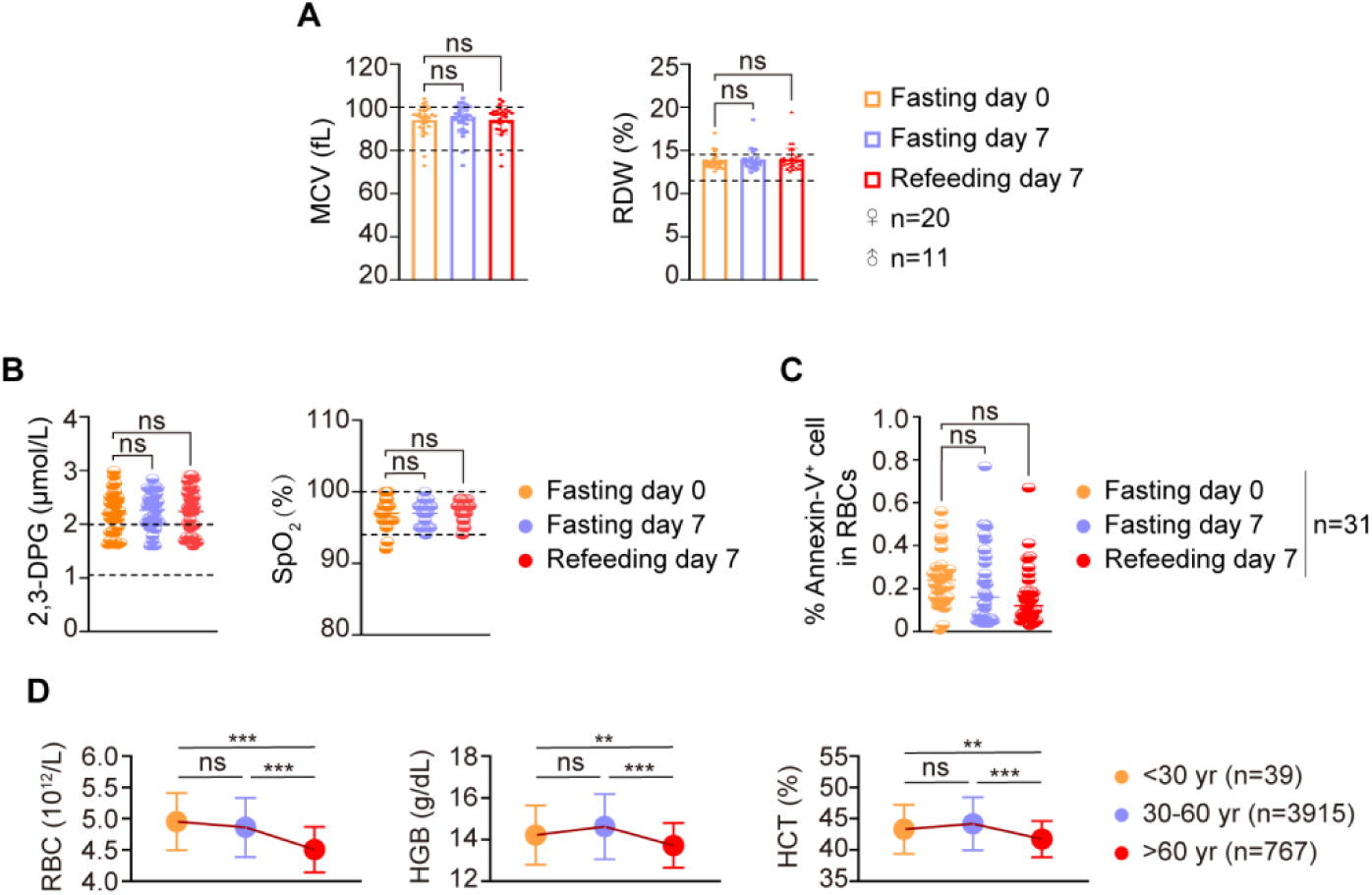
STIF rejuvenates erythropoiesis. **(A)** STIF does not alter the morphology of human peripheral red blood cells. Peripheral red blood cells were measured by routine blood count. **(B)** Left, 2,3-diphosphoglycerate (2,3-DPG) concentration of peripheral red blood cells measured by ELISA (n=31), Right, oxygen saturation determined by finger clip blood oxygen analyzer (n=23). **(C)** STIF does not alter the apoptosis level of human peripheral red blood cells. The apoptosis level of red blood cells detected by flow cytometry. **(D)** Red blood cell count declines with increasing age. Peripheral blood cells were detected by routine counting.

**Figure S2.**
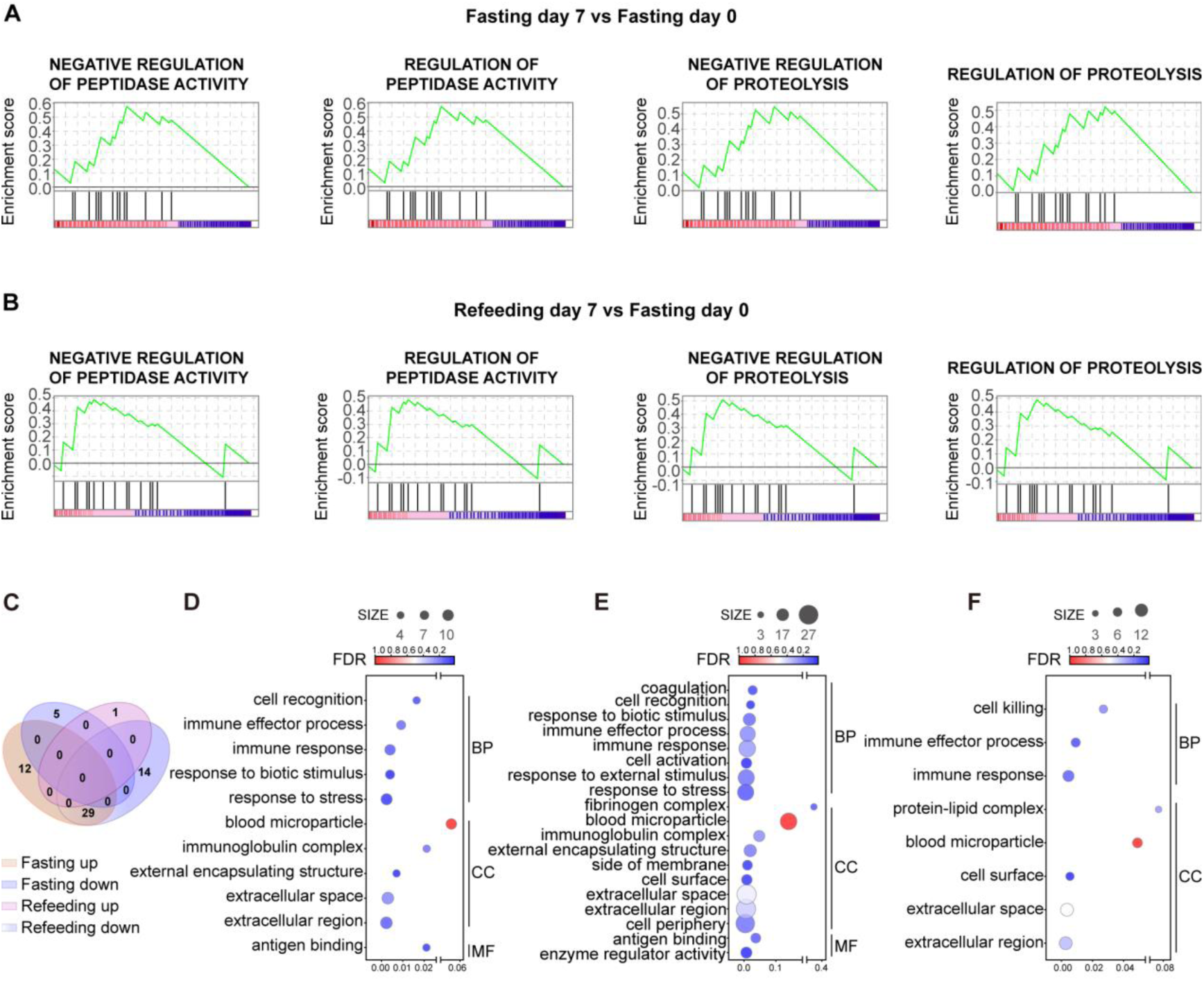
Upregulation of negative regulation of proteolysis. **(A-B)** Representative GSEA plots of proteolysis functions. **(C)** Venn diagram depicting the intersection of DEPs of two comparison groups (Fasting day 7 versus Fasting day 0 and Refeeding day 7 verses Fasting day 0). **(D-F)** Key enriched GOBP pathways identified with three types of expression patterns: upregulated at day 7 and maintained a high level of expression after refeeding (D), upregulated at day 7 but returned to normal after refeeding (E), and downregulated after refeeding (F). BP: Biological Process; CC: Cellular Component; MF: Molecular Function.

**Figure S3.**
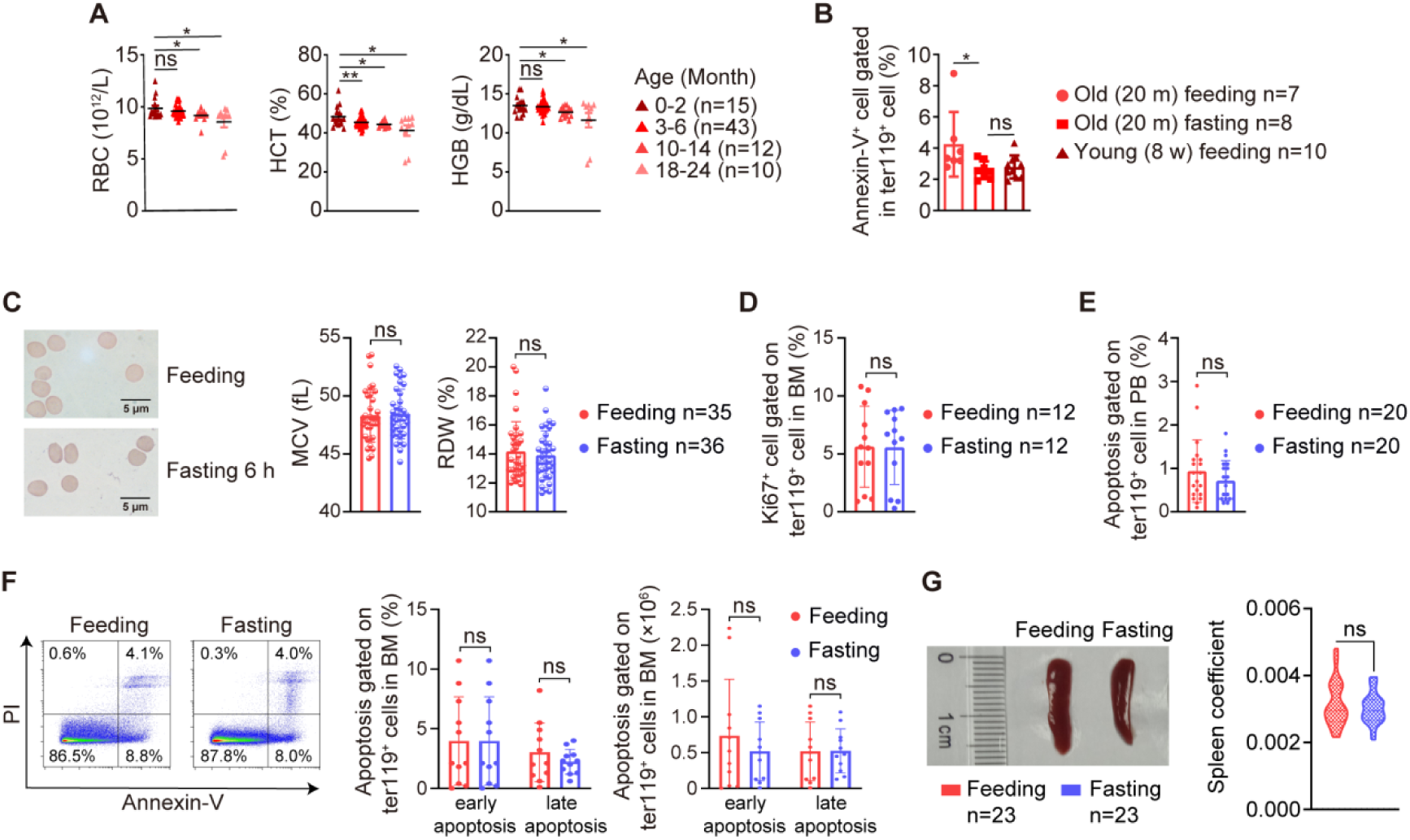
Intensive fasting impacts on bone marrow hematopoietic cells and peripheral RBCs in mice. **(A)** Fasting rejuvenates red blood cells in mice. Peripheral blood test for mice at different ages. **(B)** Apoptosis levels in mouse bone marrow red blood cells measured with Ter119 and Annexin-V flow cytometry. **(C)** Fasting does not alter the morphology of mouse red blood cells. Left, Wright-Giemsa stain; right, Mean corpuscular volume (MCV) and red cell distribution width (RDW) test. **(D)** Fasting does not alter the proliferation of bone marrow erythroid cells. The proliferation capacity of bone marrow erythroid cells was measured with Ki67 and Ter119 staining and flow cytometry. **(E-F)** Fasting does not alter apoptosis of bone marrow and peripheral erythroid cells. Apoptosis level of peripheral erythroid cells (E). Apoptosis level of bone marrow erythroid cells. The cells were measured with Annexin V/PI and Ter119 by flow cytometry (Feeding n=11, Fasting n=11) (F). **(G)** Fasting does not change the size and weight of the spleen. Left, Morphology of spleens from *Ad libitum* and 6-hour-fasting mice. Right panel, the organ coefficient (spleen/body weight) of *Ad libitum* and 6-hour-fasting mice.

**Figure S4.**
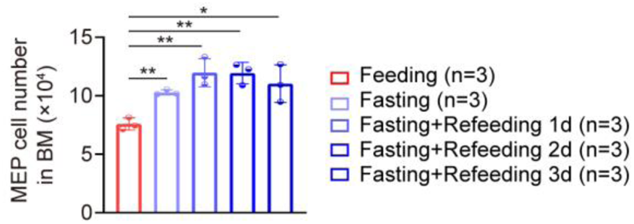
Fasting-induced upregulation of MEP proliferation is fairly sustainable in mice. Quantitation of MEP cells on a polychromatic flow cell analyzer.

**Figure S5.**
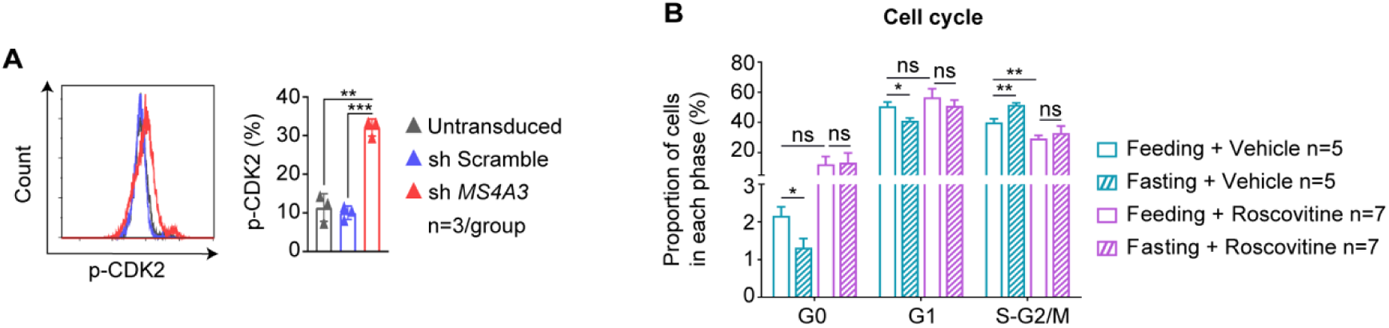
MS4A3 is located upstream of CDK2 in regulating fasting-induced erythropoiesis. **(A)** The expression of p-CDK2 decreased in MS4A3 knockdown HUDEP-2 cells. Levels of p-CDK2 were measured by flow cytometry (n=3/group). **(B)** The cell cycle of MEP cells was arrested after intraperitoneal injection of roscovitine. Ki67 and Hoechst 33342 were used to test the cell cycle of MEP.

**Figure S6.**
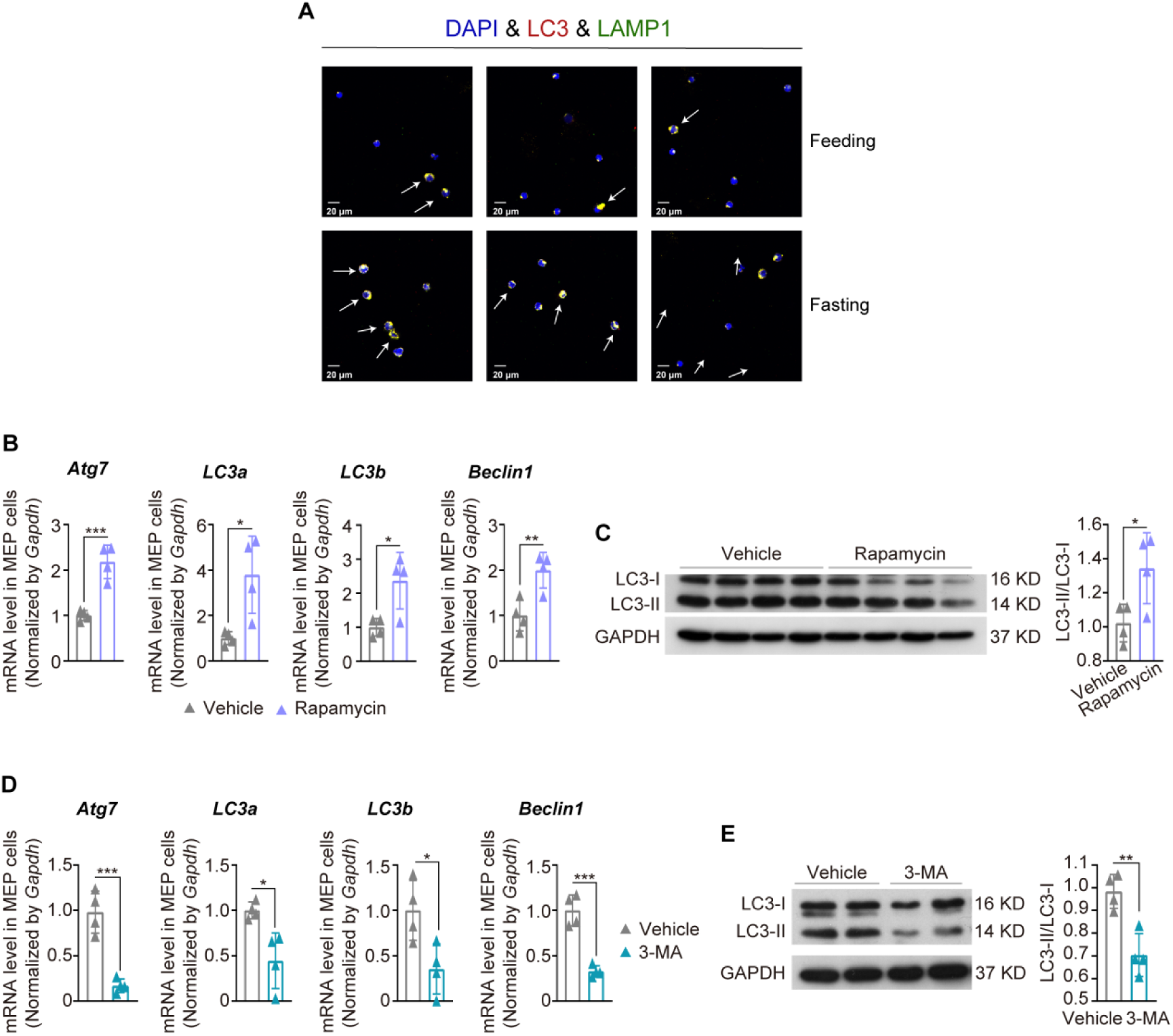
Autophagy activity is required in fasting-triggered erythropoiesis. **(A)** Fasting activates *in vivo* autophagy activity in mice. Confocal observation of autophagy markers in MEPs from *Ad libitum* and 6-hour-fasting mice by confocal microscopy. **(B-C)** Rapamycin activates *in vivo* basal autophagic activity in mice (n=4/group). Expression of *Atg 7*, *LC3a*, *LC3b* and *Beclin1* of MEP cells in mice with or without Rapamycin injection tested by Q-PCR (B). Autophagic markers of BM-MNCs from wild-type mice that received intraperitoneal injection of rapamycin were measured by Western blotting (C). **(D-E)** 3-MA inhibits *in vivo* basal autophagic activity in mice (n=4/group). Autophagic markers of BM-MNCs from wild-type mice that received intraperitoneal injection of 3-MA were measured by Q-PCR (D) and Western blotting (E).

**Figure S7.**
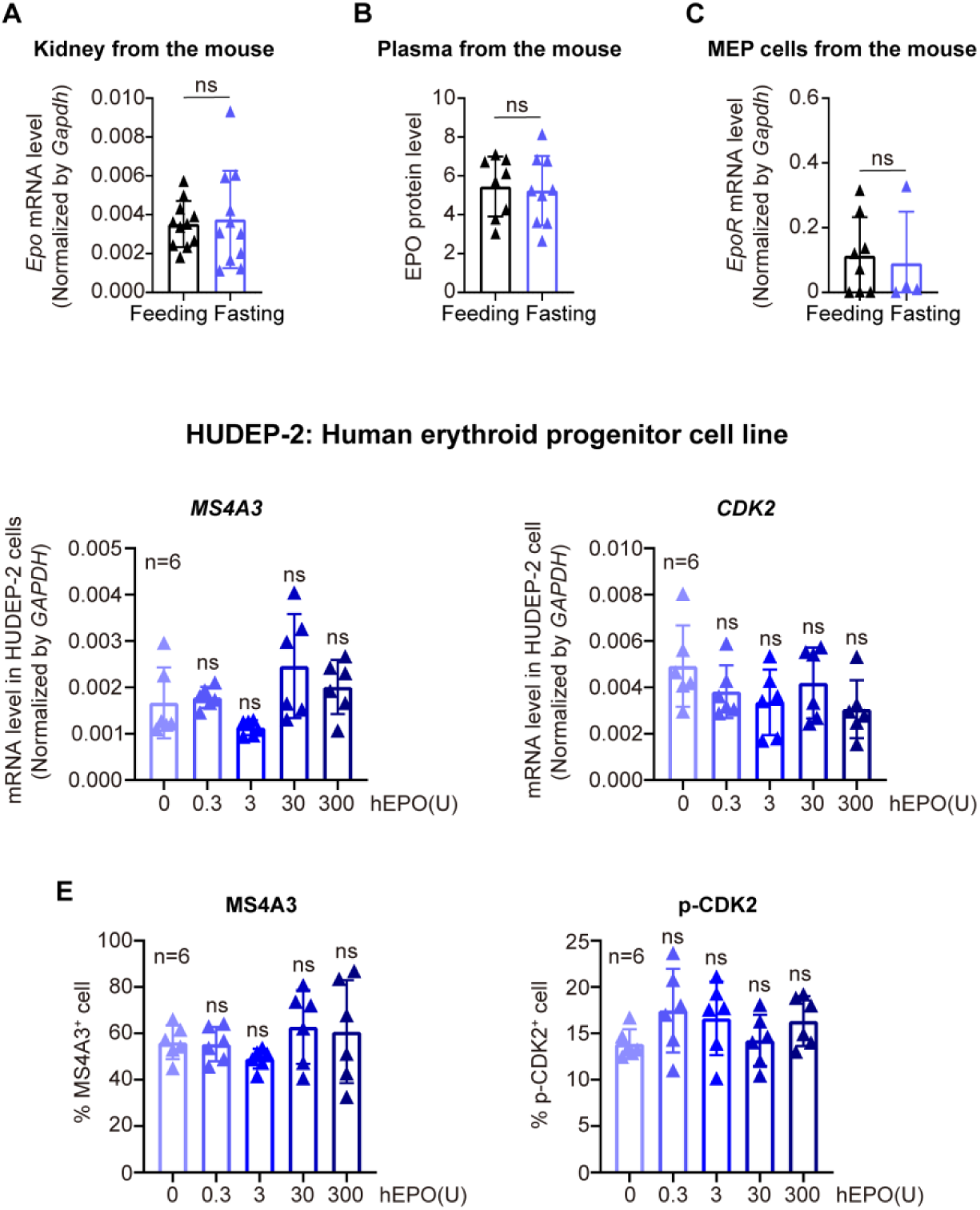
The fasting-triggered MS4A3-CDK2 module enhances erythropoiesis independent of EPO. **(A-C)** Fasting did not enhance *in vivo* EPO and EPOR expression in mice. *Epo* mRNA was measured by Q-PCR in kidney cell before and after 6 hr of fasting. n=11/group (A). EPO level in mouse plasma was measured by ELISA. Feeding n=8; Fasting n=9 (B). *EpoR* mRNA was measured by Q-PCR in MEP cells before and after 6 hr of fasting. Feeding n=8; Fasting n=4 (C). **(D-E)** An increase in EPO concentration did not influence the expression levels of *MS4A3* and *CDK2* in the HUDEP cell line. Transcript levels of *MS4A3* and *CDK2* were measured by Q-PCR (D). Protein levels of *MS4A3* and p-*CDK2* were measured with flow cytometry with antibodies against MS4A3 or CDK2 (E).

**Table S1.** Information of participating subjects in the fasting study (n=31)

| **Volunteer** | **Sex** | **Age**<br>**(year)** | **Height**<br>**(cm)** | **Weight**<br>**(kg)** | **Time of**<br>**Fasting (day)** | **Time of**<br>**Refeeding (day)** |
| --- | --- | --- | --- | --- | --- | --- |
| 1 | Female | 67-70 | 158 | 70 | 7 | 7 |
| 2 | Male | 61-65 | 170 | 72 | 7 | 7 |
| 3 | Male | 56-60 | 178 | 85 | 7 | 7 |
| 4 | Female | 56-60 | 158 | 70 | 7 | 7 |
| 5 | Female | 56-60 | 162 | 65 | 7 | 7 |
| 6 | Female | 51-55 | 158 | 50 | 7 | 7 |
| 7 | Male | 46-50 | 163 | 75 | 7 | 7 |
| 8 | Female | 46-50 | 160 | 57 | 7 | 7 |
| 9 | Male | 46-50 | 170 | 70 | 7 | 7 |
| 10 | Female | 46-50 | 167 | 51 | 7 | 7 |
| 11 | Male | 46-50 | 163 | 60 | 7 | 7 |
| 12 | Female | 44-45 | 160 | 73 | 7 | 7 |
| 13 | Female | 41-45 | 165 | 56 | 7 | 7 |
| 14 | Female | 41-45 | 162 | 60 | 7 | 7 |
| 15 | Male | 41-45 | 175 | 73 | 7 | 7 |
| 16 | Female | 36-40 | 158 | 52.5 | 7 | 7 |
| 17 | Female | 36-40 | 155 | 60 | 7 | 7 |
| 18 | Femal | 36-40 | 165 | 54 | 7 | 7 |
| 19 | Male | 36-40 | 180 | 75 | 7 | 7 |
| 20 | Female | 36-40 | 172 | 66 | 7 | 7 |
| 21 | Male | 36-40 | 160 | 62 | 7 | 7 |
| 22 | Female | 36-40 | 165 | 66 | 7 | 7 |
| 23 | Female | 31-35 | 177 | 80 | 7 | 7 |
| 24 | Female | 31-35 | 160 | 60 | 7 | 7 |
| 25 | Female | 31-35 | 180 | 82.5 | 7 | 7 |
| 26 | Male | 31-35 | 162 | 54 | 7 | 7 |
| 27 | Female | 31-35 | 177 | 72 | 7 | 7 |
| 28 | Male | 31-35 | 162 | 53 | 7 | 7 |
| 29 | Female | 31-35 | 183 | 100 | 7 | 7 |
| 30 | Male | 31-35 | 167 | 60 | 7 | 7 |
| 31 | Female | 25-30 | 169 | 81 | 7 | 7 |

**Table S2.**
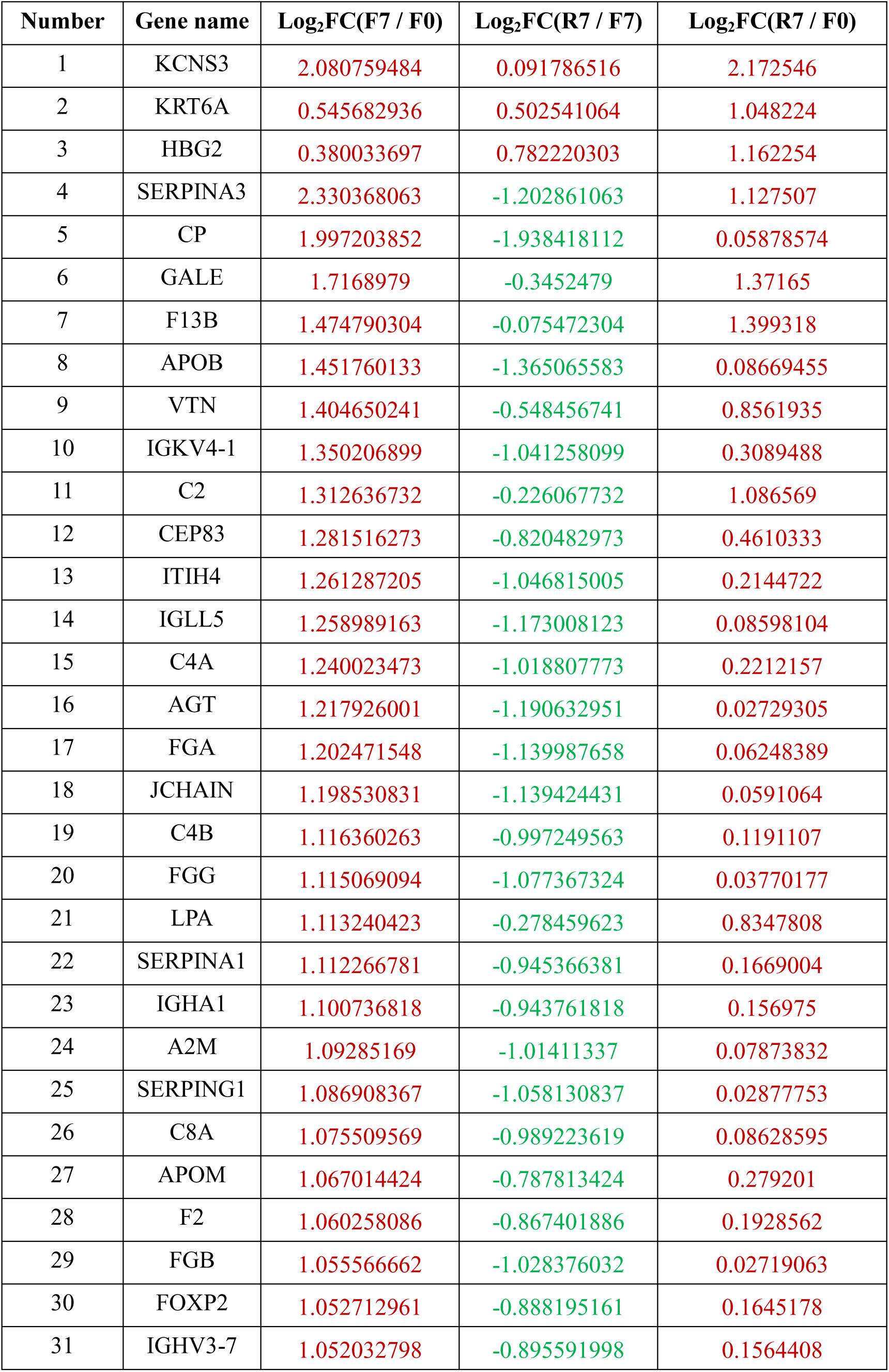
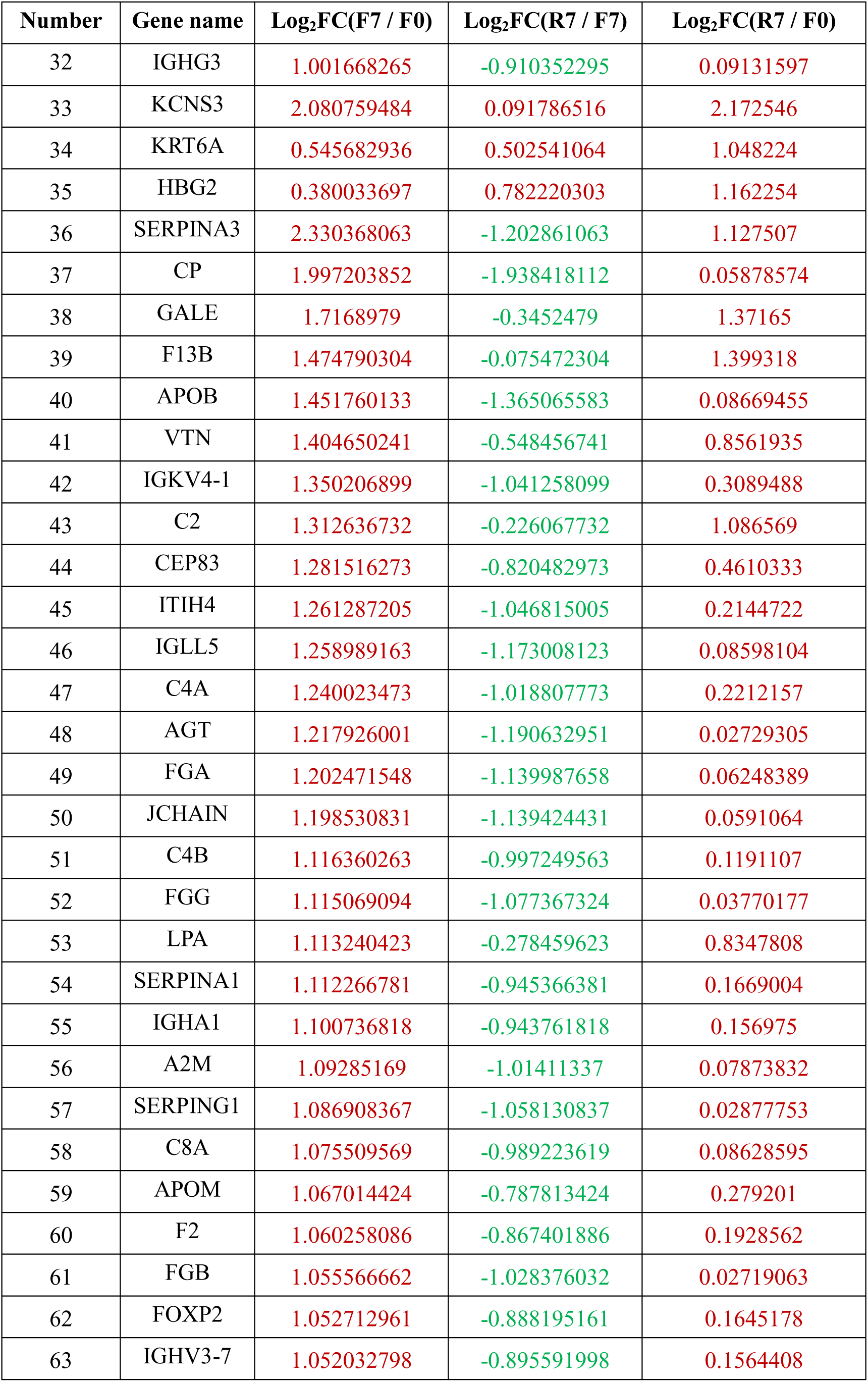
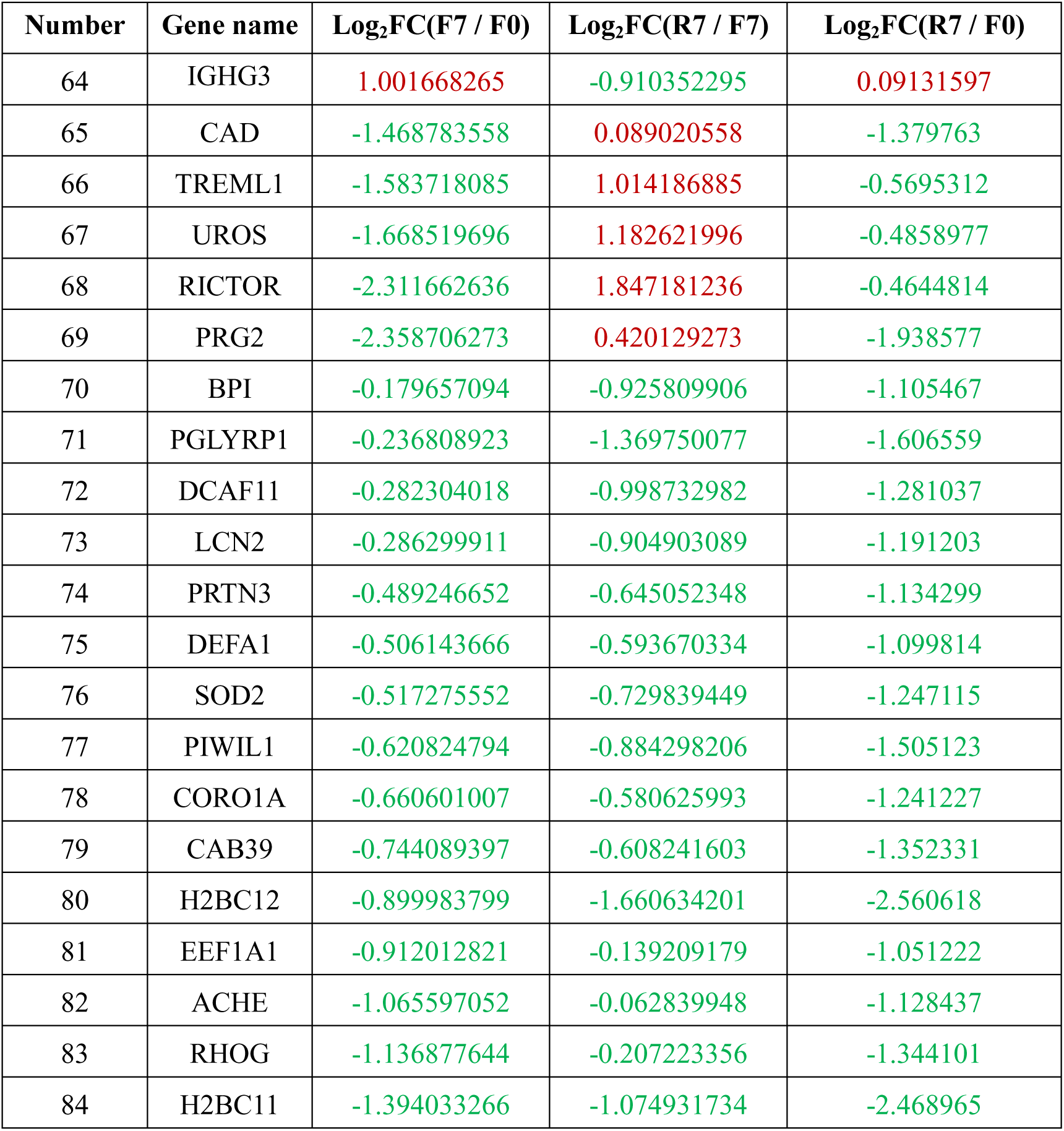
Information of 84 differentially expressed genes.

| Number | Gene name | Log<sub>2</sub>FC(F7 / F0) | Log<sub>2</sub>FC(R7 / F7) | Log<sub>2</sub>FC(R7 / F0) |
| --- | --- | --- | --- | --- |
| 1 | KCNS3 | 2.080759484 | 0.091786516 | 2.172546 |
| 2 | KRT6A | 0.545682936 | 0.502541064 | 1.048224 |
| 3 | HBG2 | 0.380033697 | 0.782220303 | 1.162254 |
| 4 | SERPINA3 | 2.330368063 | -1.202861063 | 1.127507 |
| 5 | CP | 1.997203852 | -1.938418112 | 0.05878574 |
| 6 | GALE | 1.7168979 | -0.3452479 | 1.37165 |
| 7 | F13B | 1.474790304 | -0.075472304 | 1.399318 |
| 8 | APOB | 1.451760133 | -1.365065583 | 0.08669455 |
| 9 | VTN | 1.404650241 | -0.548456741 | 0.8561935 |
| 10 | IGKV4-1 | 1.350206899 | -1.041258099 | 0.3089488 |
| 11 | C2 | 1.312636732 | -0.226067732 | 1.086569 |
| 12 | CEP83 | 1.281516273 | -0.820482973 | 0.4610333 |
| 13 | ITIH4 | 1.261287205 | -1.046815005 | 0.2144722 |
| 14 | IGLL5 | 1.258989163 | -1.173008123 | 0.08598104 |
| 15 | C4A | 1.240023473 | -1.018807773 | 0.2212157 |
| 16 | AGT | 1.217926001 | -1.190632951 | 0.02729305 |
| 17 | FGA | 1.202471548 | -1.139987658 | 0.06248389 |
| 18 | JCHAIN | 1.198530831 | -1.139424431 | 0.0591064 |
| 19 | C4B | 1.116360263 | -0.997249563 | 0.1191107 |
| 20 | FGG | 1.115069094 | -1.077367324 | 0.03770177 |
| 21 | LPA | 1.113240423 | -0.278459623 | 0.8347808 |
| 22 | SERPINA1 | 1.112266781 | -0.945366381 | 0.1669004 |
| 23 | IGHA1 | 1.100736818 | -0.943761818 | 0.156975 |
| 24 | A2M | 1.09285169 | -1.01411337 | 0.07873832 |
| 25 | SERPING1 | 1.086908367 | -1.058130837 | 0.02877753 |
| 26 | C8A | 1.075509569 | -0.989223619 | 0.08628595 |
| 27 | APOM | 1.067014424 | -0.787813424 | 0.279201 |
| 28 | F2 | 1.060258086 | -0.867401886 | 0.1928562 |
| 29 | FGB | 1.055566662 | -1.028376032 | 0.02719063 |
| 30 | FOXP2 | 1.052712961 | -0.888195161 | 0.1645178 |
| 31 | IGHV3-7 | 1.052032798 | -0.895591998 | 0.1564408 |

Continue to Table S2. Information of 84 differentially expressed genes
| Number | Gene name | Log<sub>2</sub>FC(F7 / F0) | Log<sub>2</sub>FC(R7 / F7) | Log<sub>2</sub>FC(R7 / F0) |
| --- | --- | --- | --- | --- |
| 32 | IGHG3 | 1.001668265 | -0.910352295 | 0.09131597 |
| 33 | KCNS3 | 2.080759484 | 0.091786516 | 2.172546 |
| 34 | KRT6A | 0.545682936 | 0.502541064 | 1.048224 |
| 35 | HBG2 | 0.380033697 | 0.782220303 | 1.162254 |
| 36 | SERPINA3 | 2.330368063 | -1.202861063 | 1.127507 |
| 37 | CP | 1.997203852 | -1.938418112 | 0.05878574 |
| 38 | GALE | 1.7168979 | -0.3452479 | 1.37165 |
| 39 | F13B | 1.474790304 | -0.075472304 | 1.399318 |
| 40 | APOB | 1.451760133 | -1.365065583 | 0.08669455 |
| 41 | VTN | 1.404650241 | -0.548456741 | 0.8561935 |
| 42 | IGKV4-1 | 1.350206899 | -1.041258099 | 0.3089488 |
| 43 | C2 | 1.312636732 | -0.226067732 | 1.086569 |
| 44 | CEP83 | 1.281516273 | -0.820482973 | 0.4610333 |
| 45 | ITIH4 | 1.261287205 | -1.046815005 | 0.2144722 |
| 46 | IGLL5 | 1.258989163 | -1.173008123 | 0.08598104 |
| 47 | C4A | 1.240023473 | -1.018807773 | 0.2212157 |
| 48 | AGT | 1.217926001 | -1.190632951 | 0.02729305 |
| 49 | FGA | 1.202471548 | -1.139987658 | 0.06248389 |
| 50 | JCHAIN | 1.198530831 | -1.139424431 | 0.0591064 |
| 51 | C4B | 1.116360263 | -0.997249563 | 0.1191107 |
| 52 | FGG | 1.115069094 | -1.077367324 | 0.03770177 |
| 53 | LPA | 1.113240423 | -0.278459623 | 0.8347808 |
| 54 | SERPINA1 | 1.112266781 | -0.945366381 | 0.1669004 |
| 55 | IGHA1 | 1.100736818 | -0.943761818 | 0.156975 |
| 56 | A2M | 1.09285169 | -1.01411337 | 0.07873832 |
| 57 | SERPING1 | 1.086908367 | -1.058130837 | 0.02877753 |
| 58 | C8A | 1.075509569 | -0.989223619 | 0.08628595 |
| 59 | APOM | 1.067014424 | -0.787813424 | 0.279201 |
| 60 | F2 | 1.060258086 | -0.867401886 | 0.1928562 |
| 61 | FGB | 1.055566662 | -1.028376032 | 0.02719063 |
| 62 | FOXP2 | 1.052712961 | -0.888195161 | 0.1645178 |
| 63 | IGHV3-7 | 1.052032798 | -0.895591998 | 0.1564408 |
| 64 | IGHG3 | 1.001668265 | -0.910352295 | 0.09131597 |
| 65 | CAD | -1.468783558 | 0.089020558 | -1.379763 |
| 66 | TREML1 | -1.583718085 | 1.014186885 | -0.5695312 |
| 67 | UROS | -1.668519696 | 1.182621996 | -0.4858977 |
| 68 | RICTOR | -2.311662636 | 1.847181236 | -0.4644814 |
| 69 | PRG2 | -2.358706273 | 0.420129273 | -1.938577 |
| 70 | BPI | -0.179657094 | -0.925809906 | -1.105467 |
| 71 | PGLYRP1 | -0.236808923 | -1.369750077 | -1.606559 |
| 72 | DCAF11 | -0.282304018 | -0.998732982 | -1.281037 |
| 73 | LCN2 | -0.286299911 | -0.904903089 | -1.191203 |
| 74 | PRTN3 | -0.489246652 | -0.645052348 | -1.134299 |
| 75 | DEFA1 | -0.506143666 | -0.593670334 | -1.099814 |
| 76 | SOD2 | -0.517275552 | -0.729839449 | -1.247115 |
| 77 | PIWIL1 | -0.620824794 | -0.884298206 | -1.505123 |
| 78 | CORO1A | -0.660601007 | -0.580625993 | -1.241227 |
| 79 | CAB39 | -0.744089397 | -0.608241603 | -1.352331 |
| 80 | H2BC12 | -0.899983799 | -1.660634201 | -2.560618 |
| 81 | EEF1A1 | -0.912012821 | -0.139209179 | -1.051222 |
| 82 | ACHE | -1.065597052 | -0.062839948 | -1.128437 |
| 83 | RHOG | -1.136877644 | -0.207223356 | -1.344101 |
| 84 | H2BC11 | -1.394033266 | -1.074931734 | -2.468965 |

**Table S3.** 25 key pathways identified in GSEA.

| Number | Name | NES<br>(F7 vs F0) | NES<br>(R7 vs F0) | SIZE |
| --- | --- | --- | --- | --- |
| 1 | adaptive immune response | 2.4780169 | 1.1696836 | 24 |
| 2 | complement activation | 2.3691626 | 1.288955 | 23 |
| 3 | activation of immune response | 2.3479698 | 1.3051533 | 23 |
| 4 | positive regulation of immune response | 2.3135664 | 1.272405 | 24 |
| 5 | positive regulation of immune system process | 2.3047519 | 0.8661694 | 25 |
| 6 | regulation of humoral immune response | 2.1541145 | 1.1634493 | 20 |
| 7 | regulation of immune response | 2.1390703 | 1.2111789 | 25 |
| 8 | regulation of immune effector process | 2.12183 | 1.1586803 | 20 |
| 9 | regulation of complement activation | 2.0692506 | 1.1711804 | 20 |
| 10 | b cell mediated immunity | 2.0443797 | 1.0354654 | 19 |
| 11 | lymphocyte mediated immunity | 1.9334736 | 0.5856121 | 20 |
| 12 | humoral immune response | 1.8112937 | -1.1323078 | 37 |
| 13 | regulation of immune system process | 1.7736679 | 0.5422753 | 30 |
| 14 | cell activation involved in immune response | -2.1375618 | -1.6380473 | 18 |
| 15 | proteolysis | 2.0688603 | 0.80878156 | 26 |
| 16 | regulation of proteolysis | 1.8154688 | 1.413238 | 17 |
| 17 | negative regulation of proteolysis | 1.9204016 | 1.4052136 | 16 |
| 18 | negative regulation of catalytic activity | 2.020335 | 1.2824898 | 15 |
| 19 | negative regulation of molecular function | 1.8618549 | 0.9577613 | 16 |
| 20 | regulation of peptidase activity | 1.9505483 | 1.3288373 | 15 |
| 21 | negative regulation of peptidase activity | 2.0131972 | 1.2893603 | 15 |
| 22 | negative regulation of hydrolase activity | 1.9818488 | 1.3317777 | 15 |
| 23 | myeloid leukocyte activation | -2.075185 | -1.5024601 | 18 |
| 24 | response to nitrogen compound | -2.1385796 | -1.227318 | 5 |
| 25 | organonitrogen compound biosynthetic process | -2.3904865 | -1.2844943 | 10 |

## REFERENCES

Bai M, Cao P, Lin Y, et al. Intermittent caloric restriction promotes erythroid development and ameliorates phenylhydrazine-induced anemia in mice. Front Nutr. 2022;9:892435. doi: 10.3389/fnut.2022.892435.

Betz T, Lenz M, Joanny JF, Sykes C. ATP-dependent mechanics of red blood cells. Proc Natl Acad Sci U S A. 2009;106(36):15320–15325. doi:10.1073/pnas.0904614106.

Brandhorst S, Choi IY, Wei M, et al. A periodic diet that mimics fasting promotes multi-system regeneration, enhanced cognitive performance, and healthspan. Cell Metab. 2015;22(1):86–99. doi:10.1016/j.cmet.2015.05.012.

Bruce L.J., Pan R.J., Cope D.L., Uchikawa M., Gunn R.B., Cherry R.J., Tanner M.J. Altered structure and anion transport properties of band 3 (AE1, SLC4A1) in human red cells lacking glycophorin A. J Biol Chem. 279:2414–2420 (2004).

Caffa I, Spagnolo V, Vernieri C, et al. Fasting-mimicking diet and hormone therapy induce breast cancer regression. Nature. 2020; 583(7817):620–624. doi:10.1038/s41586-020-2502-7.

Cao Y, Cai J, Zhang S, et al. Autophagy sustains hematopoiesis through targeting Notch. Stem Cells Dev. 2015;24(22):2660–2673. doi:10.1089/scd.2015.0176.

Cao Y, Zhang S, Yuan N, et al. Hierarchal autophagic divergence of hematopoietic system. J Biol Chem. 2015;290(38):23050–23063. doi:10.1074/jbc.M115.650028.

Caulier AL, Sankaran VG. Molecular and cellular mechanisms that regulate human erythropoiesis. Blood. 2021;blood.2021011044. doi:10.1182/blood.2021011044.

Chaves PH, Ashar B, Guralnik JM, Fried LP. Looking at the relationship between hemoglobin concentration and prevalent mobility difficulty in older women. Should the criteria currently used to define anemia in older people be reevaluated?. J Am Geriatr Soc. 2002;50(7):1257–1264. doi:10.1046/j.1532-5415.2002.50313.x.

Chen-Edinboro LP, Murray-Kolb LE, Simonsick EM, et al. Association Between non-iron-deficient anemia and insomnia symptoms in community-dwelling older adults: The baltimore longitudinal study of aging. J Gerontol A Biol Sci Med Sci. 2018;73(3):380–385. doi:10.1093/gerona/glw332.

Chen J, Astle CM, Harrison DE. Hematopoietic senescence is postponed and hematopoietic stem cell function is enhanced by dietary restriction. Exp Hematol. 2003;31(11):1097–1103. doi:10.1016/s0301-472x(03)00238-8.

Cheng CW, Adams GB, Perin L, et al. Prolonged fasting reduces IGF-1/PKA to promote hematopoietic-stem-cell-based regeneration and reverse immunosuppression. Cell Stem Cell. 2014;14(6):810–823. doi:10.1016/j.stem.2014.04.014.

Cheng CW, Villani V, Buono R, et al. Fasting-mimicking diet promotes Ngn3-driven β-cell regeneration to reverse diabetes. Cell. 2017;168(5):775–788.e12. doi:10.1016/j.cell.2017.01.040.

Chinami M, Yano Y, Yang X, et al. Binding of HTm4 to cyclin-dependent kinase (Cdk)-associated phosphatase (KAP).Cdk2.cyclin A complex enhances the phosphatase activity of KAP, dissociates cyclin A, and facilitates KAP dephosphorylation of Cdk2. J Biol Chem. 2005;280(17):17235–17242. doi:10.1074/jbc.M413437200.

Collins N, Han SJ, Enamorado M, et al. The bone marrow protects and optimizes immunological memory during dietary restriction. Cell. 2019;178(5):1088–1101.e15. doi:10.1016/j.cell.2019.07.049.

Culleton BF, Manns BJ, Zhang J, Tonelli M, Klarenbach S, Hemmelgarn BR. Impact of anemia on hospitalization and mortality in older adults. Blood. 2006;107(10):3841–3846. doi:10.1182/blood-2005-10-4308.

Damiani E, Adrario E, Luchetti MM, et al. Plasma free hemoglobin and microcirculatory response to fresh or old blood transfusions in sepsis. PLoS One. 2015;10(5):e0122655. doi:10.1371/journal.pone.0122655.

Danecek P, Bonfield JK, Liddle J, et al. Twelve years of SAMtools and BCFtools. Gigascience. 2021;10(2):giab008. doi:10.1093/gigascience/giab008.

de Cabo R, Mattson MP. Effects of Intermittent Fasting on Health, Aging, and Disease. N Engl J Med. 2019;381(26):2541–2551. doi:10.1056/NEJMra1905136.

Denny SD, Kuchibhatla MN, Cohen HJ. Impact of anemia on mortality, cognition, and function in community-dwelling elderly. Am J Med. 2006;119(4):327–334. doi:10.1016/j.amjmed.2005.08.027.

Di Biase S, Lee C, Brandhorst S, et al. Fasting-mimicking diet Reduces HO-1 to promote T cell-mediated tumor cytotoxicity. Cancer Cell. 2016;30(1):136–146. doi:10.1016/j.ccell.2016.06.005.

Donato JL, Ko J, Kutok JL, et al. Human HTm4 is a hematopoietic cell cycle regulator. J Clin Invest. 2002;109(1):51–58. doi:10.1172/JCI14025.

Doudna JA. The promise and challenge of therapeutic genome editing. Nature. 2020;578(7794):229–236. doi:10.1038/s41586-020-1978-5.

Eckardt KU, Boutellier U, Kurtz A, Schopen M, Koller EA, Bauer C. Rate of erythropoietin formation in humans in response to acute hypobaric hypoxia. J Appl Physiol. 1989;66(4):1785–1788. doi:10.1152/jappl.1989.66.4.1785.

Fang Y, Gu Y, Zhao C, et al. Impact of supervised beego, a traditional Chinese water-only fasting, on thrombosis and haemostasis. BMJ Nutr Prev Health. 2021;4(1):4–17. doi:10.1136/bmjnph-2020-000183.

Fernández ÁF, Sebti S, Wei Y, et al. Disruption of the bec lin 1-BCL2 autophagy regulatory complex promotes longevity in mice. Nature. 2018;558(7708):136–140. doi:10.1038/s41586-018-0162-7.

Frangoul H, Altshuler D, Cappellini MD, et al. CRISPR-Cas9 Gene Editing for Sickle Cell Disease and β-Thalassemia. N Engl J Med. 2021;384(3):252–260. doi:10.1056/NEJMoa2031054.

Fu B, Liao J, Chen S, Li W, Wang Q, Hu J, Yang F, Hsiao S, Jiang Y, Wang L, Chen F, Zhang Y, Wang X, Li D, Liu M, Wu Y. CRISPR–Cas9-mediated gene editing of the BCL11A enhancer for pediatric β0/β0 transfusion-dependent β-thalassemia. Nature Medicine 2022 Aug 4. doi: 10.1038/s41591-022-01906-z.

Galluzzi L, Pietrocola F, Levine B, Kroemer G. Metabolic control of autophagy. Cell. 2014;159(6):1263–1276. doi:10.1016/j.cell.2014.11.006.

Geng G, Liu J, Xu C, et al. Receptor-mediated mitophagy regulates EPO production and protects against renal anemia. Elife. 2021;10:e64480. doi:10.7554/eLife.64480.

Ge RL, Witkowski S, Zhang Y, et al. Determinants of erythropoietin release in response to short-term hypobaric hypoxia. J Appl Physiol. 2002;92(6):2361–2367. doi:10.1152/japplphysiol.00684.2001.

Han K, Singh K, Rodman MJ, et al. Fasting-induced FOXO4 blunts human CD4^+^ T helper cell responsiveness. Nat Metab. 2021;3(3):318–326. doi:10.1038/s42255-021-00356-0.

Hong CH, Falvey C, Harris TB, et al. Anemia and risk of dementia in older adults: findings from the Health ABC study [published correction appears in Neurology. 2013 Sep 3;81(10):939]. Neurology. 2013;81(6):528–533. doi:10.1212/WNL.0b013e31829e701d.

Jaiswal S. Clonal hematopoiesis and nonhematologic disorders. Blood. 2020;136(14): 1606–1614. doi: 10.1182/blood.2019000989.

Jaiswal S, Ebert BL. Clonal hematopoiesis in human aging and disease. Science. 2019;366(6465):eaan4673. doi: 10.1126/science.aan4673.

Jordan S, Tung N, Casanova-Acebes M, et al. Dietary intake regulates the circulating inflammatory monocyte pool. Cell. 2019;178(5):1102–1114.e17. doi:10.1016/j.cell.2019.07.050.

Kassebaum NJ, Jasrasaria R, Naghavi M, Wulf SK, Johns N, Lozano R, Regan M, Weatherall D, Chou DP, Eisele TP, Flaxman SR, Pullan RL, Brooker SJ, Murray CJL. A systematic analysis of global anemia burden from 1990 to 2010. Blood. 2014;123(5):615–24. doi: 10.1182/blood-2013-06-508325.

Kassebaum NJ. The Global burden of anemia. Hematol Oncol Clin North Am. 2016;30(2):247–308. doi: 10.1016/j.hoc.2015.11.002.

Kim D, Paggi JM, Park C, Bennett C, Salzberg SL. Graph-based genome alignment and genotyping with HISAT2 and HISAT-genotype. Nat Biotechnol. 2019;37(8):907–915. doi:10.1038/s41587-019-0201-4.

Komatsu M, Waguri S, Ueno T, et al. Impairment of starvation-induced and constitutive autophagy in Atg7-deficient mice. J Cell Biol. 2005;169(3):425–434. doi:10.1083/jcb.200412022.

Kundu M, Lindsten T, Yang CY, et al. Ulk1 plays a critical role in the autophagic clearance of mitochondria and ribosomes during reticulocyte maturation. Blood. 2008;112(4):1493–1502. doi:10.1182/blood-2008-02-137398.

Kurita R, Suda N, Sudo K, et al. Establishment of immortalized human erythroid progenitor cell lines able to produce enucleated red blood cells. PLoS One. 2013;8(3):e59890. doi:10.1371/journal.pone.0059890.

Kutok JL, Yang X, Folkerth RD, et al. The cell cycle associated protein, HTm4, is expressed in differentiating cells of the hematopoietic and central nervous system in mice. J Mol Histol. 2005;36(1-2):77–87. doi:10.1007/s10735-004-3913-8.

Liao Y, Smyth GK, Shi W. FeatureCounts: an efficient general purpose program for assigning sequence reads to genomic features. Bioinformatics. 2014;30(7):923–930. doi:10.1093/bioinformatics/btt656.

Li-Harms X, Milasta S, Lynch J, et al. Mito-protective autophagy is impaired in erythroid cells of aged mtDNA-mutator mice. Blood. 2015;125(1):162–174. doi:10.1182/blood-2014-07-586396.

Longo VD, Di Tano M, Mattson MP & Guidi N. Intermittent and periodic fasting, longevity and disease. Nature Aging. 2021; 1:47–59. doi:10.1038/s43587-020-00013-3.

Love MI, Huber W, Anders S. Moderated estimation of fold change and dispersion for RNA-seq data with DESeq2. Genome Biol. 2014;15(12):550. doi:10.1186/s13059-014-0550-8.

Lu Z, Xie J, Wu G, et al. Fasting selectively blocks development of acute lymphoblastic leukemia via leptin-receptor upregulation. Nat Med. 2017;23(1):79–90. doi:10.1038/nm.4252.

MacDonald R. Red cell 2,3-diphosphoglycerate and oxygen affinity. Anaesthesia. 1977;32(6):544–553. doi:10.1111/j.1365-2044.1977.tb10002.x.

Messaoudi I, Warner J, Fischer M, et al. Delay of T cell senescence by caloric restriction in aged long-lived nonhuman primates. Proc Natl Acad Sci U S A. 2006;103 (51):19448–19453. doi:10.1073/pnas.0606661103.

Mizushima N, Yamamoto A, Matsui M, Yoshimori T, Ohsumi Y. In vivo analysis of autophagy in response to nutrient starvation using transgenic mice expressing a fluorescent autophagosome marker. Mol Biol Cell. 2004;15(3):1101–1111. doi:10.1091/mbc.e03-09-0704.

Mortensen M, Ferguson DJ, Edelmann M, et al. Loss of autophagy in erythroid cells leads to defective removal of mitochondria and severe anemia in vivo. Proc Natl Acad Sci U S A. 2010;107(2):832–837. doi:10.1073/pnas.0913170107.

Mortensen M, Soilleux EJ, Djordjevic G, et al. The autophagy protein Atg7 is essential for hematopoietic stem cell maintenance. J Exp Med. 2011;208(3):455–467. doi:10.1084/jem.20101145.

Ogawa S. Clonal hematopoiesis in acquired aplastic anemia. Blood. 2016;128(3): 337–347. doi:10.1182/blood-2016-01-636381.

Onder G, Penninx BW, Cesari M, et al. Anemia is associated with depression in older adults: results from the InCHIANTI study. J Gerontol A Biol Sci Med Sci. 2005;60(9):1168–1172. doi:10.1093/gerona/60.9.1168.

Penninx BW, Pahor M, Cesari M, et al. Anemia is associated with disability and decreased physical performance and muscle strength in the elderly. J Am Geriatr Soc. 2004;52(5):719–724. doi:10.1111/j.1532-5415.2004.52208.x.

Pietrocola F, Pol J, Vacchelli E, et al. Caloric restriction mimetics enhance anticancer immunosurveillance. Cancer Cell. 2016;30(1):147–160. doi:10.1016/j.ccell.2016.05.016.

Poole J. Red cell antigens on band 3 and glycophorin A. Blood Rev. 2000;14(1):31–43. doi:10.1054/blre.1999.0124.

Qian J, Fang Y, Yuan N, et al. Innate immune remodeling by short-term intensive fasting. Aging Cell. 2021;20(11):e13507. doi:10.1111/acel.13507.

Robach P, Fulla Y, Westerterp KR, Richalet JP. Comparative response of EPO and soluble transferrin receptor at high altitude. Med Sci Sports Exerc. 2004;36(9):1493–1492. doi:10.1249/01.mss.0000139889.56481.e0.

Sandoval H, Thiagarajan P, Dasgupta SK, et al. Essential role for Nix in autophagic maturation of erythroid cells. Nature. 2008;454(7201):232–235. doi:10.1038/nature07006.

Schuster SJ, Badiavas EV, Costa-Giomi P, Weinmann R, Erslev AJ, Caro J. Stimulation of erythropoietin gene transcription during hypoxia and cobalt exposure. Blood. 1989;73(1):13–16. Doi:10.1182/blood.V73.1.13.13

Schweers RL, Zhang J, Randall MS, et al. NIX is required for programmed mitochondrial clearance during reticulocyte maturation. Proc Natl Acad Sci U S A. 2007;104(49):19500–19505. doi:10.1073/pnas.0708818104.

Shnitsar V., Li J., Li X., Calmettes C., Basu A., Casey J.R., Moraes T.F., Reithmeier R.A. A substrate access tunnel in the cytosolic domain is not an essential feature of the solute carrier 4 (SLC4) family of bicarbonate transporters J Biol Chem. 2013; 288:33848–33860.

Stauder R, Valent P, Theurl I. Anemia at older age: etiologies, clinical implications, and management. Blood. 2018;131(5):505–514. doi:10.1182/blood-2017-07-746446.

Subramanian A, Tamayo P, Mootha VK, et al. Gene set enrichment analysis: a knowledge-based approach for interpreting genome-wide expression profiles. Proc Natl Acad Sci U S A. 2005;102(43):15545–15550. doi:10.1073/pnas.0506580102.

Tang D, Tao S, Chen Z, et al. Dietary restriction improves repopulation but impairs lymphoid differentiation capacity of hematopoietic stem cells in early aging. J Exp Med. 2016;213(4):535–553. doi:10.1084/jem.20151100.

Ulgherait M, Midoun AM, Park SJ, et al. Circadian autophagy drives iTRF-mediated longevity. Nature. 2021;598(7880):353–358. doi:10.1038/s41586-021-03934-0.

Watts D, Gaete D, Rodriguez D, et al. Hypoxia Pathway Proteins are Master Regulators of Erythropoiesis. Int J Mol Sci. 2020; 21(21):8131. doi:10.3390/ijms21218131.

Wu Y, Zeng J, Roscoe BP, et al. Highly efficient therapeutic gene editing of human hematopoietic stem cells. Nat Med. 2019;25(5):776–783.doi:10.1038/s41591-019-0401-y.

Xu C, He J, Wang H, et al. Single-cell transcriptomic analysis identifies an immune-prone population in erythroid precursors during human ontogenesis. Nat Immunol. 2022 Jun 27. doi: 10.1038/s41590-022-01245-8.

Young M.T., Beckmann R., Toye A.M., Tanner M.J. Red-cell glycophorin A-band 3 interactions associated with the movement of band 3 to the cell surface. Biochem J. 2000; 350:53–60.

Zhang H, Shen Z, Hogan B, Barakat AI, Misbah C. ATP Release by Red Blood Cells under Flow: Model and Simulations. Biophys J. 2018;115(11):2218–2229. doi:10.1016/j.bpj.2018.09.033.

Zhang J, Randall MS, Loyd MR, et al. Mitochondrial clearance is regulated by Atg7-dependent and -independent mechanisms during reticulocyte maturation. Blood. 2009;114(1):157–164. doi:10.1182/blood-2008-04-151639.

